# SAHDAI-XAI Subarachnoid Hemorrhage Detection Artificial Intelligence- eXplainable AI: Testing explainability in SAH Imaging Data and AI Modeling

**DOI:** 10.1101/2025.09.26.25336583

**Authors:** Sarah Morgan, Saif Salman, Jordan Walker, W David Freeman

## Abstract

**Introduction:** Subarachnoid hemorrhage (SAH) is a life-threatening and crucial neurological emergency. SAHDAI-XAI (Subarachnoid Hemorrhage Detection Artificial Intelligence) is a cloud-based machine learning model created as a binary positive and negative classifier to detect SAH bleeding seen in any of eight potential hemorrhage spaces. It aims to address the lack of transparency in AI-based detection of subarachnoid hemorrhage.

**Methods:** This project is divided into two phases, integrating Auto-ML and BLAST, combining the statistical assessment of hemorrhage detection accuracy using a low-code approach with the simultaneous colour-based visualization of bleeding areas to enhance transparency. In phase 1, an AutoML model was trained on Google Cloud Vertex AI after preprocessing. The Model completed four runs, progressively increasing the dataset size. The dataset is split into 80% for training, 10% for validation, and 10% for testing, with explainability (XRAI) applied to the testing images. We started with 20 non-contrast head CT images followed by 40, 200, and then 300 images, and in each AutoML run, the dataset was equivalently divided into one half manually labeled as positive for hemorrhage and the other half labeled as negative controls. The fourth AutoML evaluated the model’s ability to differentiate between a hemorrhage and other pathologies, such as tumors and calcifications. In phase 2, the goal is to increase explainability by visualizing predictive image features and showing the detection of hemorrhage locations using the Brain Lesion Analysis and Segmentation Tool for Computed Tomography (BLAST). This model segments and quantifies four different hemorrhage and edema locations.

**Results:** In phase one, the first two AutoML runs demonstrated 100% average precision due to the small data size. In the third run, the average precision was 97.9% after increasing the dataset size, and one false negative (FN) image was detected. In the fourth round, after evaluating the model’s differentiation abilities, the average precision rate dropped to 94.4%. This round demonstrated two false positive (FP) images from the testing deck. After extensive preprocessing using the BLAST model public Python code in the second phase, topographic images of the bleeding were demonstrated with different outcomes. Some accurately cover a significant percentage of the bleeding, whereas others do not.

**Conclusion:** The SAHDAI-XAI model is a new image-based SAH explainable AI model that shows enhanced transparency for AI hemorrhage detection in daily clinical life and aims to overcome AI’s untransparent nature and accelerate time to diagnosis, thereby helping decrease the mortality rates.^6^ BLAST model utilization facilitates a better understanding of AI outcomes and supports the creation of visually demonstrated XAI in SAH detection and predicting hemorrhage coverage. The goal is to resolve AI’s hidden black-box aspect, making ML model outcomes increasingly transparent and explainable. **Keywords:** SAH, explainable AI, GCP, AutoML, BLAST, black-box.

## 1 Introduction and Literature Review

### 1.1 Subarachnoid Hemorrhage: An Overview

Subarachnoid hemorrhage (SAH) is a critical neurological emergency defined by bleeding in the subarachnoid space that surrounds the brain and spinal cord. Causes can be split into two main types: traumatic or nontraumatic. Approximately 85% of nontraumatic SAH cases are attributed to a ruptured intracranial aneurysm. Traumatic SAH is the most prevalent form of intracranial bleeding and is frequently seen in computed tomography (CT) scans of traumatic brain injuries.^1^ The global incidence of aneurysmal subarachnoid hemorrhage (aSAH) is approximately 6.1 cases per 100,000 people, with a visible increase in women over the age of 55 years.^2^ Among all emergency department visits, headaches account for 2% of cases, with SAH occurring in 1-3%. Patients face a 15% chance of death before reaching the hospital, a 25% chance of dying in the first 24 hours, and 40% don’t survive the first 30 days.^1,2^ When diagnosed early and treated accurately, the mortality can be significantly lowered to approximately 18%.^6^ Risk factors leading to aneurysmal rupture and thus leading to SAH include hypertension, smoking, alcohol abuse, sympathomimetic medications, and aneurysms exceeding 7mm. The most common presentation of SAH is a sudden and severe headache. However, this kind of bleeding affects not only the central nervous system but also other system of the body. Patients who survive may encounter a wide range of complications ranging from ECG abnormalities and cardiac complications^3^ and a large proportion 33-66% develop delayed cerebral ischemia (DCI) that leads to delayed neuronal injury presumably caused by cortical spreading depolarizations with or without delayed arterial narrowing (“vasospasm”) of the large cerebral arteries.^4^ Since clinical examination alone cannot differentiate between an ischemic stroke and an intracranial hemorrhage^5^, the standard approach is to perform a non-contrast head CT (NCCT). This imaging method serves not only for diagnosing but also for excluding other issues such as cancer, brain bleeding, or abscesses. When the NCCT results are inconclusive, the following step is to perform a lumbar puncture to look for red blood cells (RBC) or xanthochromia in the cerebrospinal fluid (CSF). Magnetic resonance imaging (MRI) and computed tomography angiography (CTA) are further diagnostic techniques for SAH identification. Ensuring the airway is secured and the hemodynamics are stable is the first and most important step after confirming a SAH diagnosis. Reversing anticoagulation and lowering the patient’s systolic blood pressure are the next actions to reduce the risk of recurrent aneurysmal rupture. To prevent further recurrence, treatment should include analgesia, antiemetics, and stool softeners to reduce pain, nausea, and Valsalva.^6^ Additionally, studies indicate that the calcium channel blocker nimodipine is essential for preventing one of the main complications of SAH, cerebral vasospasm (CVS), thereby reducing mortality and the risk of rebleeding.^7^ Generally, it is essential to focus on timely and accurate detection, coupled with fast intervention, to improve patient outcomes and reduce mortality rates.^8^

### 1.2 SAH Diagnostic and Outcome Predictions

In cases of SAH, medical professionals apply various scoring and grading systems to evaluate neurological status. Commonly used Classification scores include the Hunt and Hess classification, which asses the severity of SAH based on clinical neurological signs, and the World Federation of Neurosurgical Surgeons (WFNS) score, which combines the Glasgow Coma Scale (GSC) with evaluation of motor deficits. Figure 1 shows another essential grading scale, the Modified Fisher Scale (mFS)^9^. It is a radiographic system that evaluates the amount and location of bleeding and assesses the risk of DCI.^10^. Research has shown that SAH’s blood volume is a key prognostic factor. Despite this, mFS has a limited prognostic value because of inadequate metric definitions and its poor ability to differentiate between hemorrhage grades.^11^ Moreover, a comparative study indicates that models that automatically quantify total blood volume (TBV) and then separately assess cisternal, intraventricular (IVH), and intraparenchymal (IPH) blood volumes offer superior predictive accuracy compared to mFS.^11^ One alternative approach to estimating the total volume of subarachnoid hemorrhage (SAHV) on NCCT is to apply a mathematical ellipsoid formula to measure the amount of blood in the basal cisternal compartments. When tested against manual computerized segmentation, this method revealed comparable results with no significant difference in accuracy. Both methods showed a correlation between higher SAHV, which was linked to worse outcomes, and an elevated risk of DCI.^12^ A subsequent study developed an enhanced volumetric scoring system SAH (eSAH) score for aneurysmal subarachnoid hemorrhage. This scoring system is designed to anticipate the risk of DCI, assess disability through the modified Rankin Scale (mRS), and predict mortality rates. Attempting to facilitate an improved patient screening for SAH and expedite management processes, the eSAH score combines factors like GCS, age, and SAHV.^13^

**Figure 1:**
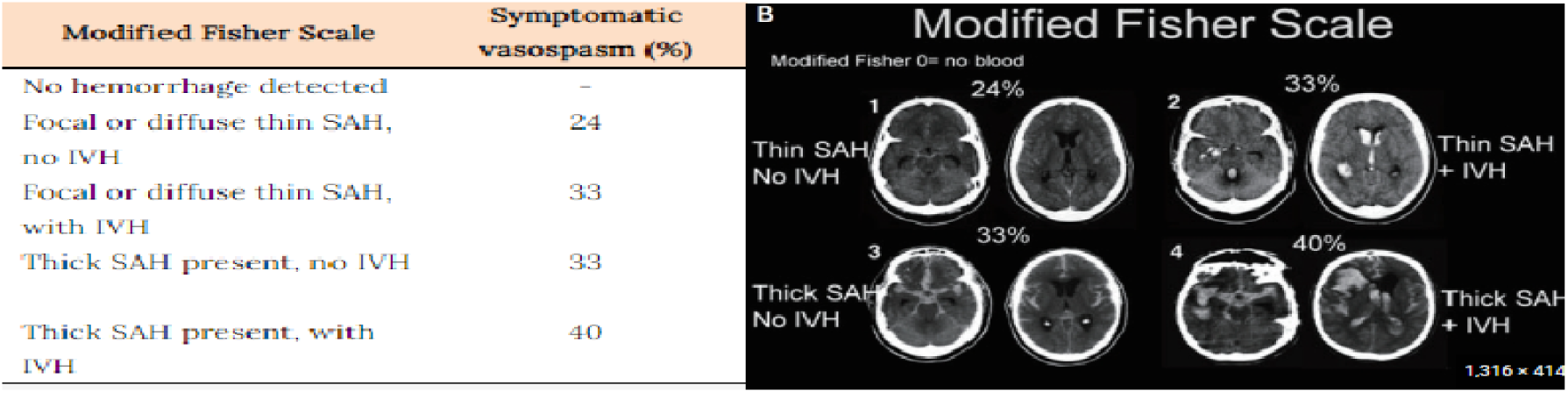
modified Fisher scale (mFS) Source: Transcranial Doppler Monitoring in Subarachnoid Hemorrhage. Adapted from Park, Kim and Ko ^9^ et al.

### 1.3 Artificial Intelligence Overview and Role in Medicine

Artificial intelligence (AI) was developed to perform tasks like those done by humans, such as object identification, pattern recognition, and problem-solving. Machine learning (ML) is a section of AI that enables systems to learn from data and identify the connection between features and patterns. ML can show and predict diagnoses and choose recommended treatment options in the medical field by analyzing given inputs like medical records or imaging without requiring explicit preprogramming.^14,15^ In the past few years, AI and ML tools have shown the ability to improve medical diagnostic accuracy and efficiency, and they have often even outperformed clinicians.^16^ Subarachnoid hemorrhage is a fatal condition in which timely intervention is crucial to prevent worsening outcomes. Multiple studies have demonstrated that AI can significantly enhance diagnostic accuracy, improve outcome prediction, and reduce the time between imaging and treatment (Figure 2).^17^ Techniques that include total blood volume calculation and NCCT have also been shown to contribute to these improvements.^18^

**Figure 2:**
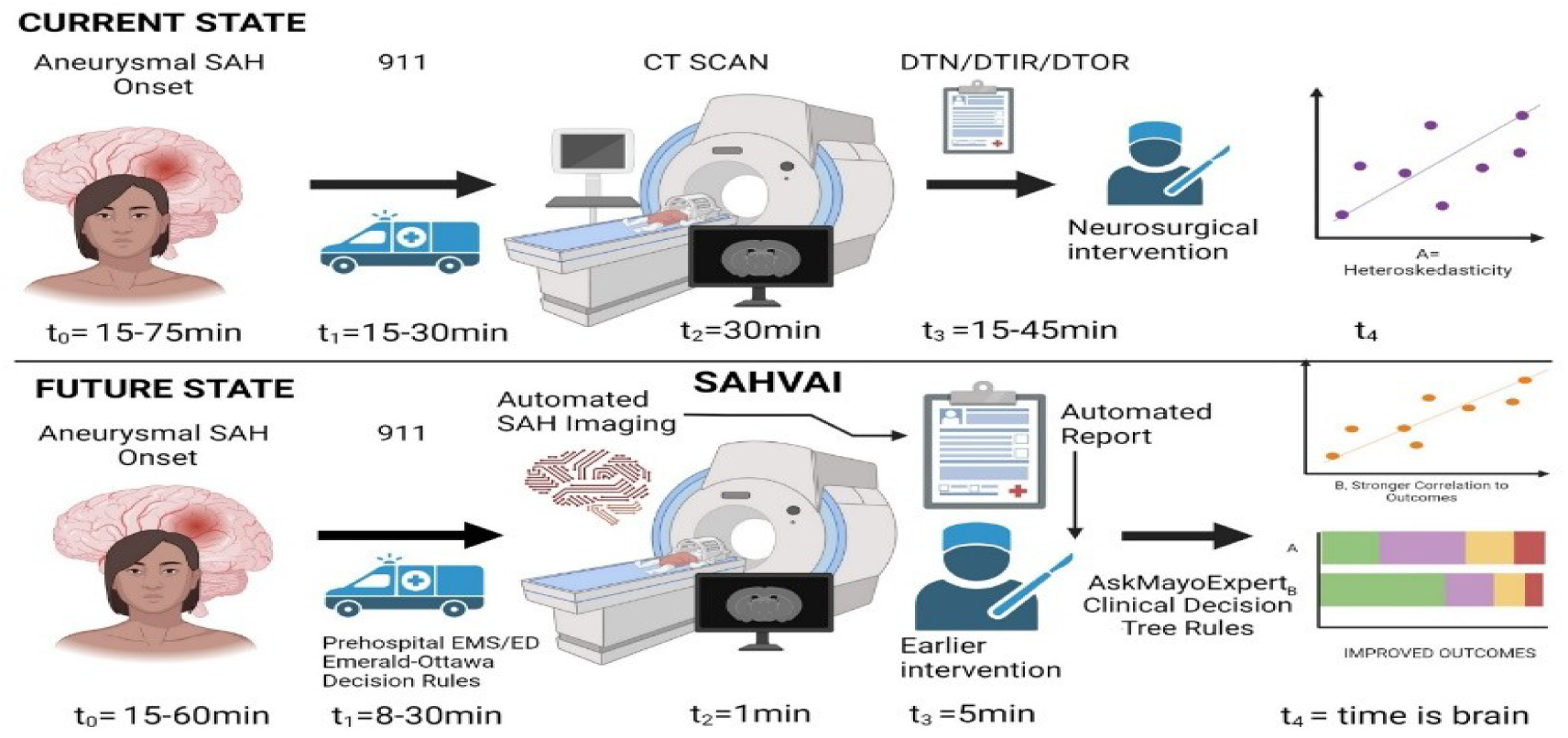
SAHVAI (SAH Volumetric AI) expediting management: Schematic illustration depicting how AI can accelerate image analysis and, thus, effectively reduce the time between imaging and intervention by BioRender.com)

#### 2.3.2 SAH AI methods: Medical Application example: SAHVAI model

##### (An Automated Subarachnoid Hemorrhage Volumetric Artificial Intelligence model)

In 2023, a systematic review was conducted using resources from the National Library of Medicine (NLM) to explore AI and ML brain imaging methods utilization for subarachnoid hemorrhage (SAH).^18^ The goal was to recognize current limitations and explore how AI could automate the measurement of SAH blood volume. Based on the review, a new technique called SAH Volumetric AI (SAHVAI-3D) was developed. A specialized AI-ML method is used in this approach to generate SAHVAI values alongside a detailed 3D map that visualizes the SAH blood distribution of SAH blood. This method is utilized to evaluate neurological outcomes by measuring the modified Rankin Scale (mRS) and analyzing areas with the highest concentration of blood, examining their association with outcomes such as vasospasm (VSP) and delayed cerebral ischemia (DCI). Parallel to that, a manual model was developed to pre-segment the NCCT scan, thus enhancing the model’s reliability. It was designed to target eight specific regions, including the five cisternal spaces (suprasellar, Sylvian, prepontine, peri mesencephalic, and interhemispheric cisterns) and, additionally, intraparenchymal, interventricular, and sulcal/gyral areas were analyzed. The AI-ML-based method demonstrated greater precision and effectiveness than the manual technique, detecting and quantifying the blood volume in 6.7 seconds compared to the manual method, which took approximately one hour, making the automated method 536.6 times faster. Furthermore, it was evident that the SAHVAI brain map and volume recorded at admission significantly correlate with subsequent neurological outcomes. These findings indicate that the AI model offers exceptional accuracy and speed in diagnosing and predicting prognosis for SAH.^19^

### 1.4 Artificial Intelligence Challenges and opportunities

In the last decade, significant advancements in AI and machine learning have become increasingly influential across various fields, achieving the capability of solving overly complex problems. Once trained, computers can answer questions based on their prior training. However, explaining AI outputs has become challenging in developing deep neural network systems and advanced deep learning (DL) models. As a result, traditional AI systems are considered “black boxes” as they provide answers without explanation.^20^ In many scenarios, especially within the medical field where critical and time-sensitive decisions have to be made, it is essential for the predicted outputs to be transparent, justifiable, and reliable. Moreover, it is crucial to offer clear explanations to prevent biases that could lead to severe consequences.^20,21^ Additionally, ensuring the safety, effectiveness, and fairness of AI models is a responsibility that extends beyond ethical considerations and must be fulfilled by legal standards. Starting in 2024, new regulations have influenced AI usage and require companies to be transparent about their training and testing processes for medical decision support systems. An important factor that has been considered is, for example, prohibiting discrimination in medical AI algorithms based on race, color, national origin, gender, age, or disability.^22^ An explainable (XAI) solution has been developed to address the challenge of understanding black boxes. Ultimately, XAI aims to overcome issues of transparency that have been restricting AI integration into critical domains. The core goal of explainable AI is to use different techniques to improve the interpretability, trustworthiness, and overall reliability of AI systems. This aspect is essential for reducing the potential consequences of misdiagnosis and patient outcomes.^23,24^

While numerous AI-based methods for SAH like the above mentioned SAHVAI model and other methods have shown promise in accelerating diagnostic processes, increasing accuracy and supporting outcomes prediction and risk assesment^54,55,56^, a major limitation still persists because of the lack transparency in how these models generate their results and clinicians are unaware of how the model comes at its conclusions. This is precisely where explainable AI (XAI) comes to its conclusions.

### 1.5 AI Explainability (XAI) methods

An AI model’s prediction becomes understandable and explainable when it shows how the input data leads to the output in a way that makes sense to the user. This involves highlighting the most relevant factors that led to the outcome, making the connection between input and prediction easy for the user to follow. The growing number of eXplainable AI (XAI) approaches makes assessing their strengths, weaknesses, and suitability for different domains difficult.^25^ XAI methods vary widely in their approach to model interpretation. For example, Local Interpretable Model-Agnostic Explanation (LIME) explains a technique called explanation by simplification. It simplifies complex model predictions by treating the model as a black box and using a more understandable surrogate model to mimic the behavior around a specific input. In contrast, Shapley Additive Explanations (SHAP) offers explanations based on feature relevance by quantifying the contribution of each variable to a model’s prediction.^26^ On the other hand, Explanation with Ranked Area Integrals (XRAI) enhances interpretability by focusing on image regions rather than individual pixels. It presents a rough map of critical areas and then refines them by aggregating pixel-level attributions, resulting in clearer and more comprehensible explanations.^27^

### 1.6 Cloud Computing

Cloud computing allows users to access and utilize many computing resources like servers, storage, and applications via the Internet. These resources are available and released without users having to manage the hardware or engage deeply with the service provider.^28^ Low-code development minimizes manual coding and allows people with limited programming expertise to create and use software often integrated with cloud technologies.^29^ This combination of cost efficiency and the convenience of remote access poses great advantages and challenges, including ensuring data security and privacy and achieving service reliability must be taken into consideration. Despite these issues, cloud computing is overcoming conventional limitations, offering disaster recovery solutions to minimize problems, and supporting advanced technologies such as AI. This promotes ongoing progress and creates future opportunities.^28,29^ In the medical field, it is apparent that cloud computing has revolutionized healthcare by offering effective solutions for handling large datasets. It can manage complex computing tasks, including image segmentation and feature extraction, with the utilization of numerous virtual machines to perform operations more rapidly and efficiently.^30^ Moreover, it supports data analytics and machine learning, encouraging better connections among healthcare professionals via collaborative platforms. This results in more personalized medical treatments and improved general patient outcomes.^31^

### 1.7 GCP and AutoML

Google Cloud Platform (GCP) is a selection of cloud-based tools and services offered by Google. Despite its relatively recent introduction, it plays a major role in the cloud market by providing various services that utilize Google’s infrastructure.^32^ It includes simplified AI services, such as AutoML, designed to make ML accessible to users without specialized expertise. AutoML automates the entire process of building custom ML models, reducing manual effort and improving efficiency. It is effective for tasks such as image classification and object detection. It can also manage all data processing steps, from preprocessing and feature engineering to model selection and hyperparameter optimization. These facilities allow users to create customized AI projects with minimal effort.^33,34^

### 1.8 Quantification and Segmentation of Brain Imaging

Brain image quantification involves detecting and quantifying specific brain structures and features, like blood volume measurement. Segmentation, on the other hand, emphasizes brain imaging scans partitioning into separate regions according to different anatomical structures to present a detailed analysis. Both quantification and segmentation play a crucial role in brain CT imaging, which enhances diagnostic efficiency, disease progression monitoring, and patient treatment.^35^ The Brain Lesion Analysis and Segmentation Tool for Computed Tomography (BLAST) model focuses on voxel-wise multi-class-segmentation and quantification of traumatic brain injury lesions of head CT using a neural network. It serves as an illustration to underline the importance of automated quantification and segmentation in the medical field. A study led by Monteiro et al. mainly concentrates on the analysis of head CT scans of traumatic brain injury (TBI) patients. It emphasizes the significance of this approach, given that TBI is the leading cause of death in young adults. In this study, the presented model calculates the total blood volume of each lesion and segments four main areas, including intraparenchymal hemorrhage (IPH), extra-axial hemorrhage (EAH), perilesional edema, and intraventricular hemorrhage (IVH). This aims to improve brain lesion detection, quantification, and segmentation efficiency and accuracy.^36^

### 1.9 Application of AI Methods in SAH diagnosis : Current applications and remaining gaps

Recent research in SAH has applied AI to support patient outcome predictions, identify treatment risks and personalize therapeutic approaches.^55,56^ AI has also demonstrated the ability to identify prognostic factors associated with poor outcomes, including rebleeding, DCI and vasospasm.^58,59,60^ Moreover, AI has shown that it can expedite the detection of SAH on CT scans, aiming to reduce diagnostic delays.^54,55,56^ Especially, explainability methods like SHAP have increased the transparency of AI models by demonstrating the influence of individual clinical features, such as age, GCS and mFS on model outputs. By breaking down each prediction into its contributing variables, SHAP allows for an understanding of how specific features contribute to the decision making and thereby increasing clinicianś confidence.^55^ However, despite these advances, there remains a lack of research focusing only on explainability in SAH detection based purely on imaging features. Addressing this research gap could further enhance AI explainability and strengthen the trust in AI model adoption for SAH detection among clinicians.^61^

## 3. Scientific Hypothesis and Aims

This thesis aims to demonstrate the importance of XAI in medical AI applications, specifically focusing on neurocritical cases like subarachnoid hemorrhage. Our goal is to illustrate how XAI models can enhance trust among healthcare professionals and acquire legal permission by proving their reliability and transparency for the presented model, particularly focusing on aSAH. Since the use of AI in aSAH detection continues to raise questions about how a machine truly knows what it knows, the role of XAI in ensuring transparency and trust becomes increasingly important. We also want to show how XAI can allow clinicians to verify and validate AI predictions, advance medical knowledge by revealing previously unnoticed imaging features, and promote the ethical use of AI in healthcare. Therefore, we hypothesize that explanations provided by AI models will help clinicians detect systematically biased AI models. To test this hypothesis, this project will explore different strategies by dividing the project into two phases. The first step is running Automated Machine Learning (AutoML) on Google Cloud Platform (GCP) to evaluate AI explainability for SAH detection, and the second stage is directed at visualizing the outputs. We will test our hypothesis by focusing on three specific aims:

***<u>Aim1</u>***: Investigate the GCP Vertex AI AutoML SAHDAI model approach for SAH image detection evaluating sensitivity, specificity, precision, and accuracy, all integrated within the GCP. Transparency should be evaluated by measuring the percentage of cases in which the model correctly identifies the presence or absence of SAH on CCT scans.

***Aim2***: Present explainable topographic visualization of SAH bleeding, focusing primarily on the imaging components of SAH and recognize aggressive bleeding patterns while assessing the use of the BLAST-CT model for SAH detection.

***<u>Aim3</u>***: Combine BLAST-SAHDAI-XAI aiming to enhance transparency by clearly visualizing the precise locations of the bleeding, thereby enhancing the modeĺs reliability and aligning with clinical expectations. Compare the AI generated segmentations to manually annotated bleeding regions in CCT scans by human experts, relying solely on visual interpretation and imaging characteristics of SAH patterns to assess concordance and support explainability. Additionally, determine the statistical correlation between AI model outputs and human expert annotations through quantitative analysis.

## 4 Materials and Methods

### 4.1 Phase 1

#### 4.1.1 General Approach

Our project for detecting SAH with artificial intelligence includes several key phases. The process started with a manual detection phase, where potential hemorrhage areas within any of the eight spaces were identified. Each image slice was manually segmented, and any suspected bleeding areas were highlighted using the ITK-SNAP application.^37^ In the next step, each image was annotated as either positive or negative for bleeding in an Excel sheet. The annotations were based on previously completed annotations, created through the combined efforts of clinical experts such as neuroradiologists, neurologists and neurosurgeons. These annotated images were then uploaded to Google Cloud Platform (GCP). On GCP, a machine learning model was created and trained using AutoML, automatically selecting the optimal training settings to distinguish between slices with bleeding and those without bleeding. Finally, the XRAI option was selected before initiating the training process. This is designed to show explainability for the images after the training phase.^38^ Moreover, the ML model processes images with 256 ^3^ RGB (Red, Green, Blue) values per pixel, displaying every pixel with three values for each RGB. This results in approximately 16.777.216 million shades per pixel, allowing the AI to detect and analyze structures beyond the human eye’s capabilities. ^39^

#### 4.1.2 Data Collection and Pre-Processing

For data collection, a large dataset of positive SAH NCCT scans from the Mayo Clinic in Jacksonville, Florida, between 2012 and 2022, along with several negative scans showing no bleed, was downloaded. The patient demographics varied in age and gender, and the clinical presentation included a range of mFS degrees, GCS scores, and Hint and Hess scores upon admission. Additionally, outcomes related to vasospasm, DCI, and mRS scores were included to support the detection of SAH in NCCT scans using AI. Initially, a total of 320 non-contrast CT scans (NCCT) were randomly selected and used, including 150 cases of SAH, 150 normal clear scans, and 20 scans representing different pathologies like calcified brain tumors. There are no indications of inclusion or exclusion criteria, as the selected CCT images are anonymized before being used and uploaded to the GCP, making it impossible to trace them back to individual patients. These anonymized images were categorized into two annotation groups, one being hemorrhage (positive) and the other no-hemorrhage (negative), with the original dataset being saved in Digital Imaging and Communication in Medicine (DICOM) format. In the following step, it was necessary to convert the data to PNG format to upload the images to GCP and use AutoML. After uploading the PNG images on GCP, each slice was manually labeled as positive or negative based on how it was previously manually annotated.

#### 4.1.3 Model Training and Evaluation

Four rounds of AutoML were done in this experiment to demonstrate explainability in the bleeding detection process. The training sample size started at 20 images and was increased to 300. Primarily, in the first round (V1),10 positive and 10 negative SAH scan slices from the uploaded dataset were randomly selected for analysis and labeled accordingly. Subsequently, AutoML on Google Cloud Platform (GCP) was used to process the data by selecting the most suitable machine learning model for training and validation. The entire process was anticipated to take 8 hours. However, it was completed in less than 2 hours. During this time, the 20 images were divided into three sets (16:2:2): the training set (80% or 16 Images), which aims to minimize loss values and get the best analysis possible; the validation set (10% or two images), which assess the learning rate and selects hyperparameters to enhance training; and the testing set (10% or two images), which evaluates the model’s performance, determines its ability to identify hemorrhage presence accurately and most importantly is the set where explainability is shown.

In the second round (V2), 20 image slices were added to expand the dataset by adding further analysis of explainability. The images were divided into (32:4:4). In the third round, 100 positive and 100 negative images were added and divided into (160:20:20), demonstrating an average precision of 97.7% and presenting one false positive (FP) image. Finally, in the fourth round (V4), 150 positive and 150 negative images were selected to enhance explainability and challenge our model further. The negative control set included 20 FP images of meningiomas and other hyperdense structures, such as basal ganglia calcifications, to assess whether our trained model could detect and correctly differentiate between bleedings and hyperdense structures.

### 4.2 Phase 2

#### 4.2.1 BLAST-SAHDAI-XAI versus Human

In the second round, the main target was to show explainability using different visualization settings. The primary priority was demonstrating clarification and justification for a model’s bleeding detection localization. Initially, different preprocessing methods, such as finding the right code and the appropriate settings for explainability, were assessed to achieve the best visualization possible. Subsequently, a public pre-trained Python-based model code called BLAST on GitHub was selected.^40^ This model focuses on quantifying and detecting hemorrhagic brain lesions and edema in traumatic brain injuries (TBI). Additionally, the code has a brain lesion segmentation ability for lesion for head CTs, and it can also calculate the volumes of these hemorrhagic lesions. This further enhances the model’s suitability for our next step in showing explainability for SAH detection and calculating the hemorrhage blood volume in the segmented area, which correlates with the outcome predictions.^36^ Since the project’s second phase was strictly descriptive, the focus was on assessing the BLAST model’s ability to detect and accurately segment SAH bleeding. This was then compared to a human assessment of the bleeding based on the ground-truth SAH patterns demonstrated in Figure 3.

**Figure 3:**
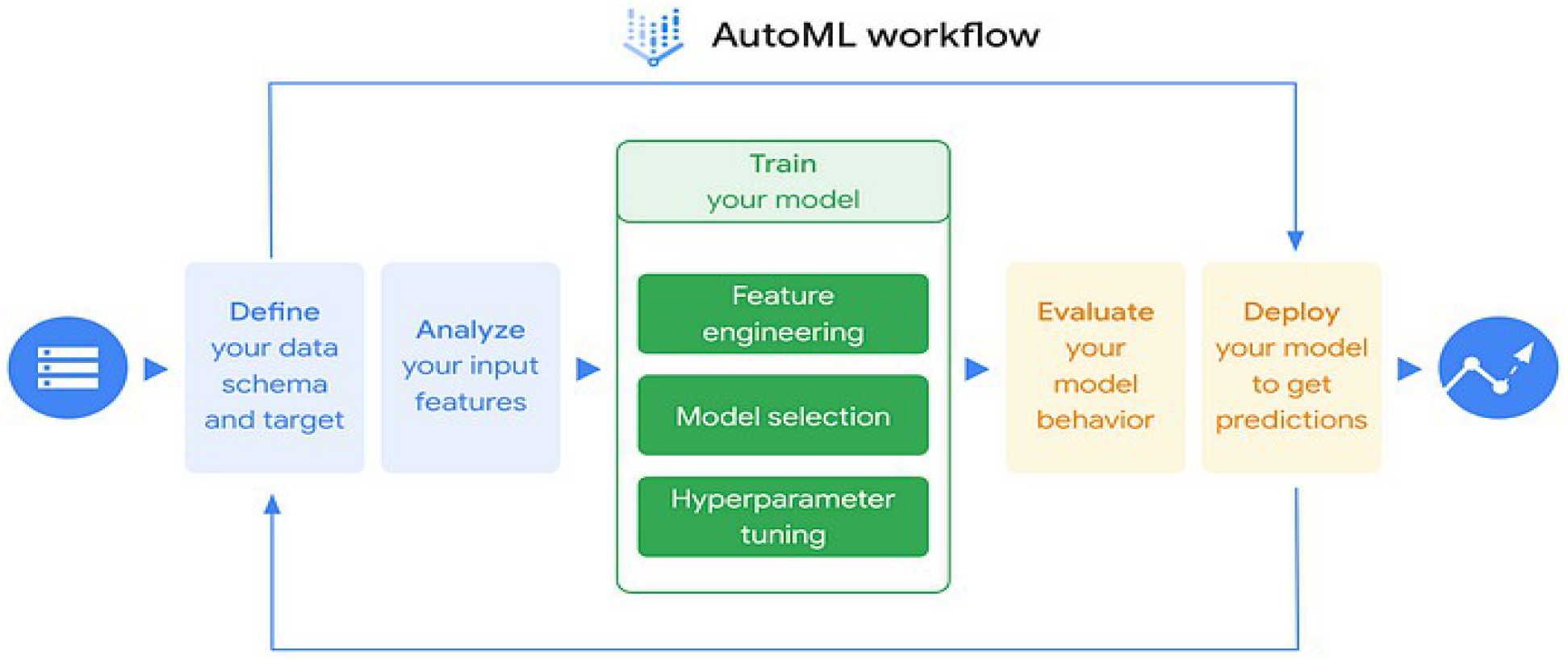
Illustration of how GCP and AutoML simplify and automate the machine learning pipeline^57^

#### 4.2.2 Phase 2 Data Preprocessing

The same 150 positive SAH patients’ images utilized in the first phase were used again in the project’s second phase. However, to be able to run the BLAST Python code on the cases, it was necessary to convert the files to a Neuroimaging Informatics Technology Initiative (NIfTI) format. After the files’ conversion process, only 146 patient cases were accessible. Primarily, the BLAST model public Python code from GitHub was utilized to help execute a code script customized for the SAHDAI-XAI-BLAST model with the assistance of an AI engineer. After that, the model was developed in Python, and interaction was established with the BLAST-CT pre-trained model weights via a command line interface (CLI). The Python code script was employed to generate predictions for areas where SAH is detected, and masked overlay images were additionally created to compare the model’s prediction to the ground truth source images.

#### 4.2.3 BLAST-XAI versus human: Descriptive Output Visualization Assessment

The project’s second phase focuses on assessing and comparing the model’s ability to detect and predict the bleeding areas topographically. The Python code was modified to demonstrate the images’ source, prediction, and overlay for each slice of the 146 patients in a linear sequence within one unified image. This was created to facilitate an easier comparison of the ground truth NCCT to the model’s predictions, thereby presenting an easier anatomical comparison and visual explainability. To estimate the model’s SAH detection ability qualitatively and descriptively, the color coverage of the masked images was assessed based on the coverage of the eight known potential hemorrhage spaces. The BLAST-CT model segments four categories, IVH, IPH, edema, and EAH, each in a different color. For each case, a BLAST-XAI model versus human ground analysis was performed, and every case was subjectively categorized into one of three categories based on the approximation of the hemorrhage detection, especially considering the mask predictions and color coverage. The three categories are determined based on the following criteria: <u>Good</u>: 60% to 100% of the hemorrhage detected and colored ➔ The midpoint is 80% <u>Moderate</u>: 30% to 60% of hemorrhage detected and colored ➔ The midpoint is 45% <u>Bad:</u> 0% to 30% of hemorrhage detected and colored ➔ The midpoint is 15%

#### 4.2.4 BLAST numerical prediction

Another important feature of the BLAST-CT model is hemorrhage blood volume quantification in different regions of head CT images. To assess the model’s performance, the Dice score for the segmentation accuracy was calculated for each patient slice,^41^ comparing the segmented regions of BLAST-CT to the manually segmented ground truth cases created by pixel-level segmentation annotations, which three physicians supervised. The code was finetuned to detect IPH, IVH, and EAH, leaving the edema detection out of the quantification as it has a non-contributary role in SAH volumetric detection. Eventually, the Dice score of the BLAST model is compared to the aSAH-specific SAHV model. Dice equation is defined as (A,B) = 2*|A ∩B| / |A| +|B|.

### 4.3 statistical methods: SAHV manual human ground truth vs

BLAST-CT eah volume output For a better performance evaluation of the BLAST-CT model, its predicted blood volumes were compared to manually calculated subarachnoid hemorrhage volumes (SAHV), which served as the ground truth. The predictions were generated using two configurations, one on a system with only a central processing units (CPU) and a single model, and the other on a graphics processing units (GPU) accelerated machine using a collection of pre-trained models. The GPU configuration offers advantages in handling large image data due to its strength while some algorithms may still perform better on a CPU depending on their structure.^62^ For comparison, three columns of data were created in Excel. The manually calculated SAHV blood volumes (ground truth), the BLAST-CT predictions from the CPU based model, and the predictions from the GPU based model. This allowed for a comparative statistical analysis between the CPU model and the ground truth, as well as between the GPU model and the ground truth.

During data preparation, a few cases had missing values in the manually calculated SAHV data. These incomplete data points were removed, which resulted in a final dataset of 84 SAH cases from initially 146 cases that were suitable for analysis. We started with a basic data analysis in Excel, including descriptive statistics such as mean, median, mode, variance, standard deviation, and 95% confidence intervals for both the CPU and GPU predictions, each compared to the SAHV ground truth values. Moreover, to evaluate the relationship between the model outputs and the ground truth, we performed regression analysis and ANOVA, which provided us with outputs like regression coefficients and residual output values. Residual volume assessment is especially important, because it reflects the differences between predicted and actual values, helping to evaluate if the BLAST-CT AI model tends to overestimate or underestimate blood volumes in different cases. Performing these comparisons allowed us to assess how closely the model’s predictions aligned with reference values calculated by clinical experts and thereby demonstrating the potential of the BLAST model for clinical integration, supported by clear statistical evidence.

## 5 Results

### 5.1 Phase 1

#### 5.1.1 Statistical Findings

Precision-recall and average precision rate percentages were demonstrated with a threshold of 0.5. Additionally, a confusion matrix was presented, showing the number of true positives (TP), true negatives (TN), false positives (FP), and false negatives (FN). In the first AutoML run (V1), the 20- slides dataset showed 100% sensitivity(recall) and precision, with the area under the curve (AUC) being 1.- No explainability could be demonstrated with a very small dataset, like 20 slides, and a 100% sensitivity without any false prediction. After trying to add 20 image slices (V2), there was no noticeable change in the performance, and the sensitivity remained at 100%. In the third run (V3), the average precision was 97.7% after increasing the dataset size. One false negative (FN) image was detected, which had been manually labeled as positive before the machine learning training, but the model detected it as negative. In the fourth round, after adding 150 positive, 130 negative, and 20 hyperdense structures manually labeled as negative, the average precision rate dropped to 94.4% and the sensitivity to 93.3%. This round demonstrated two false positive (FP) images from the testing deck, indicating that the images were manually labeled as negative because they were classified as other pathologies, such as brain tumors, but the model identified them as hemorrhages.

#### 5.1.2 Explainability and Data Interpretation

The outcome percentages on GCP’s Vertex AI AutoML are presented, showing high and low true positives and true negatives based on the clarity and extent of the bleeding in the CT image. This enhances the model’s explainability.

Focusing on the first two rounds(V1/V2), we suspect the dataset was too small to show any explainability. In the third round, with 200 images, we started seeing the first explainability demonstration with one FN result (C1). This image was manually labeled as positive, with SAH suspected in front of the pons area. Still, our SAHDAI-XAI model predicted it as negative, likely identifying a hyperdense basilar artery and not bleeding. Two FP images were conveyed in the fourth and final round, with 300 images, including 20 hyperdense structures, added to the negative control deck. In the first case (C2), the image is labeled negative because of an isodense right frontal tumor with surrounding hypodense vasogenic edema but no signs of bleeding. The model predicted this image as very high positive, likely due to confusion between tumors and bleeding. We suspect the model primarily focuses on detecting hyperdense regions within typically hypodense areas, such as CSF. In the second case (C3), the image did not include any bleeding and thus was labeled as negative. However, the model predicted it as low positive, likely due to confusion between an artifact and bleeding.

### 5.2 Phase 2

#### 5.2.1 BLAST-XAI Model Descriptive and Qualitative Performance Assessment for SAH Detection

After running the BLAST-XAI prediction code on the 146 cases, which took 2.5 minutes to 3 minutes each, every case was evaluated based on the color coverage of the hemorrhages and assigned to one of three categories. The prediction outcomes were designed to mask the source images, demonstrating an overlay to enhance the visual explainability of the outcomes.

An Excel sheet was created to evaluate every case descriptively and to assess the model’s overall performance. If the case was categorized as good, with a value between 60% and 100%, the midpoint value of 80 is noted. If the case was categorized as moderate, with a 30% and 60% range, the midpoint value 45 is entered. Finally, if the case was classified as bad with values ranging from 0% to 30%, the midpoint value 15 is documented. Subsequently, a graph was created to demonstrate the outcomes.

Finally, the overall performance of the model is evaluated, demonstrating that 36.99% of the images were documented as good, 30.82% as moderate, and 32.19% were recorded as bad. After calculating the weighted average based on the midpoint values, the model has an approximate 48.29% hemorrhage detection ability, concluding its moderate performance.

#### 5.2.2 BLAST Model Quantitative Analysis

A quantitative analysis of subarachnoid hemorrhage volume was performed using BLAST-CT on both CPU and GPU configurations, with results compared against manually calculated ground truth volumes (SAHV) measured in millilitres. The descriptive statistics for this comparison are presented in (Table 2). As mentioned in the methods section, due to missing data, the final analysis included a total of 84 SAH cases. A clear difference between the ground truth and the BLAST-CT results was observed. For example, the mean SAHV in the ground truth was 15.580 mL of blood volume while BLAST-CT measured 5.458 mL of blood volume using the CPU and 5.322 mL using the GPU (Table 2). Although both configurations underestimated the blood volume, the CPU results were slightly closer to the ground truth. For this reason, the BLAST-CT CPU model was chosen as the primary comparator for further analysis.

**Table 1:**
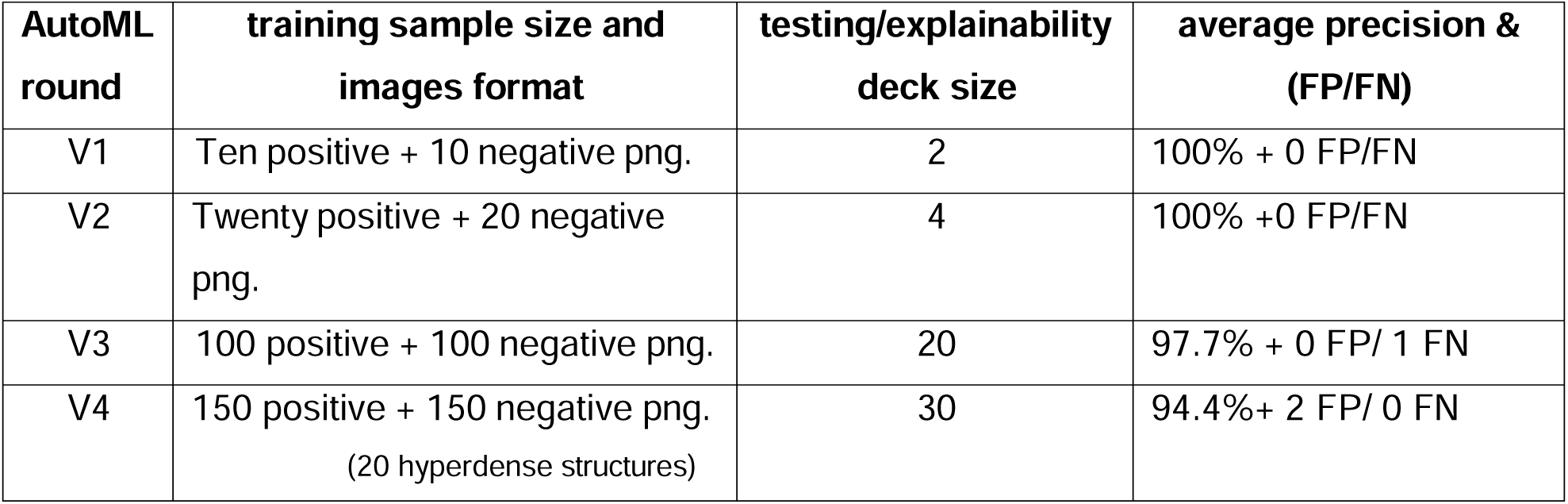
4 AutoML outcomes

| AutoML round | training sample size and images format | testing/explainability deck size | average precision & (FP/FN) |
| --- | --- | --- | --- |
| V1 | Ten positive + 10 negative png. | 2 | 100% + 0 FP/FN |
| V2 | Twenty positive + 20 negative png. | 4 | 100% +0 FP/FN |
| V3 | 100 positive + 100 negative png. | 20 | 97.7% + 0 FP/ 1 FN |
| V4 | 150 positive + 150 negative png.<br>(20 hyperdense structures) | 30 | 94.4%+ 2 FP/ 0 FN |

**Table 2:** Descriptive analysis of SAH blood volume in mL: ground truth (SAHV) vs. BLAST-CT (CPU and GPU)

|  | *SAHV (ground truth)* | *BLAST-CT (CPU)* | *BLAST-CT (GPU)* |
| --- | --- | --- | --- |
| Mean | 15.58004819 | 5.45759036 | 5.322168675 |
| Standard Error | 1.344043891 | 1.50134056 | 1.516209334 |
| Median | 13.194 | 2.09 | 0.98 |
| Mode | 0 | 0.23 | 0 |
| Standard Deviation | 12.2448226 | 13.6778634 | 13.81332443 |
| Sample Variance | 149.9356804 | 187.083948 | 190.8079318 |
| Kurtosis | 1.698078535 | 44.8540476 | 45.37267365 |
| Skewness | 1.29631493 | 6.19942913 | 6.223544728 |
| Range | 54.62 | 111.05 | 112.34 |
| Minimum | 0 | 0 | 0 |
| Maximum | 54.62 | 111.05 | 112.34 |
| Sum | 1293.144 | 452.98 | 441.74 |
| Count | 83 | 83 | 83 |
| Confidence Level (95.0%) | 2.673731454 | 2.98664463 | 3.016223365 |

The comparison between ground truth SAHV and BLAST-CT AI predicted values reveals several notable differences. On average, the AI model significantly underestimates the hemorrhage volume, as shown by the above-mentioned mean values. This underestimation is underlined further in the median values (2.09 mL in AI model vs. 13.194 mL in ground truth) and the total volume sum (452.98 mL in AI model vs. 1293.144 mL in ground truth). Moreover, despite similar standard errors, the AI output shows greater variability, as indicated by a higher standard deviation (13.678 in AI vs. 12.2448 in ground truth) and sample variance (187.08 vs. 149.93). The AI model also demonstrates a skewed and peaked distribution, with a skewness of 6.199 and kurtosis of 44.854, compared to 1.296 and 1.698 in the ground truth data. This suggests the presence of outliers with sometimes extreme overestimations. Additionally, the range of AI predictions is nearly double that of the ground truth (111.05 mL vs. 54.62 mL), confirming the inconsistency. Although both datasets share the same minimum value of 0, the AI model reaches a maximum value of 111.05 mL, which is far higher than the ground truth maximum of 54.6. These findings indicate that the BLAST-CT model has the tendency to underestimate the blood volume compared to the ground truth measurements. (Table 2)

To provide a clearer statistical overview of the summary output, a regression analysis was performed using ANOVA in Excel. (Table 3-3.2)

**Table 3:**
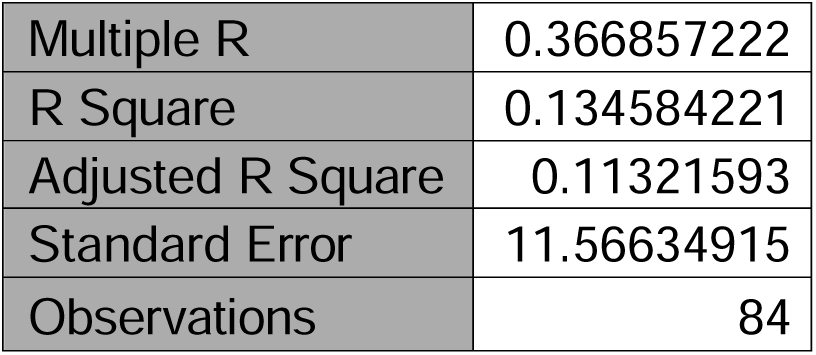
Regression statistics

|  |  |
| --- | --- |
| Multiple R | 0.366857222 |
| R Square | 0.134584221 |
| Adjusted R Square | 0.11321593 |
| Standard Error | 11.56634915 |
| Observations | 84 |

**Table 3.1:** ANOVA statistics.

|  | *df* | *SS* | *MS* | *F* | *Significance F* |
| --- | --- | --- | --- | --- | --- |
| Regression | 2 | 1685.182542 | 842.5913 | 6.29831 | 0.002868169 |
| Residual | 81 | 10836.21504 | 133.7804 |  |  |
| Total | 83 | 12521.39758 |  |  |  |

**Table 3.2:** Regression coefficients table.

|  | *Coefficients* | *Standard Error* | *t Stat* | *P-value* | *Lower 95%* | *Upper 95%* |
| --- | --- | --- | --- | --- | --- | --- |
| Intercept | 13.80004 | 1.363073263 | 10.12421 | $4.8 \times 10^{-16}$ | 11.08795041 | 16.512127 |
| CPU | -0.44811 | 0.761435001 | -0.588513 | 0.55783 | -1.963131377 | 1.0669022 |
| GPU | 0.763752 | 0.754006191 | 1.012925 | 0.31411 | -0.73648424 | 2.2639874 |

The regression ANOVA tables demonstrate that the regression model is statistically significant, with the Significance F indicating a p-value of 0.003, meaning at least one of the predictors contributes to explaining the outcome. However, the R Square value of 0.135 indicates that only 13.5% of the variability in the dependent variable is explained by the model. The low R Square and high standard error of 11.566 imply that while the model has statistical significance, its predictive accuracy is limited and that there is a lot of variation that is not described by the model. (Table 3 and 3.1)

There was also an intercept variable included in the regression analysis and two predictors CPU and GPU BLAST-CT outputs were included. The intercept, which reflects the estimated SAHV when both predictors are zero, was 13.8000 and highly significant with a p value<0.0001 indicating a stable baseline estimate. The CPU coefficient was -0.448 and the GPU coefficient was 0.763, which demonstrate minor changes in the predicted volume. However, neither one of the coefficients reached statistical significance (*p-value= 0.557* for CPU and *p-value= 0.314* for GPU), and both confidence intervals included zero, implying that these predictors did not contribute to the model’s performance. (Table 3.2 and Figure 14)

**Figure 4:**
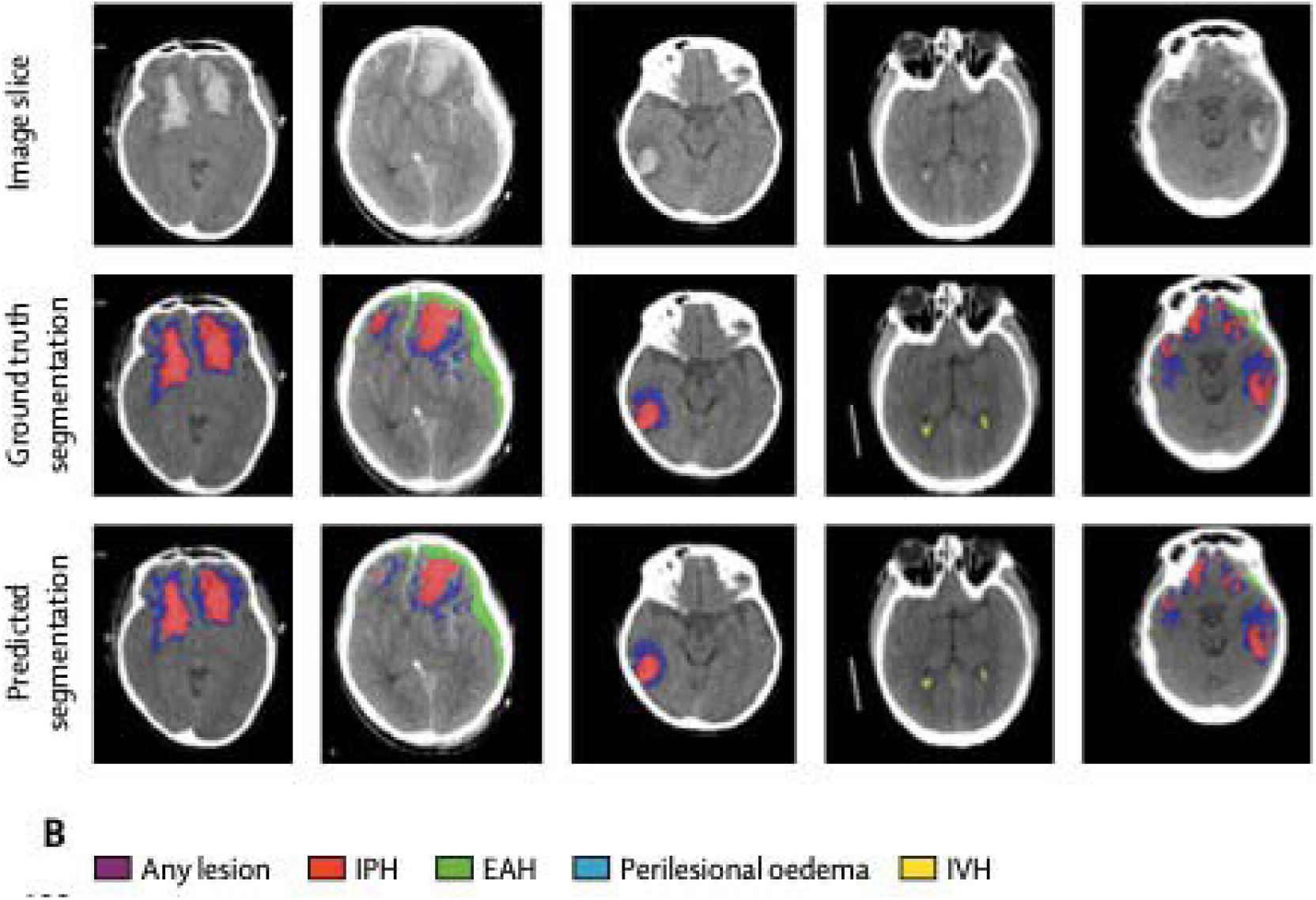
Comparison of BLAST modeĺs automated segmentation with manual ground truth segmentation^36^

**Figure 5:**
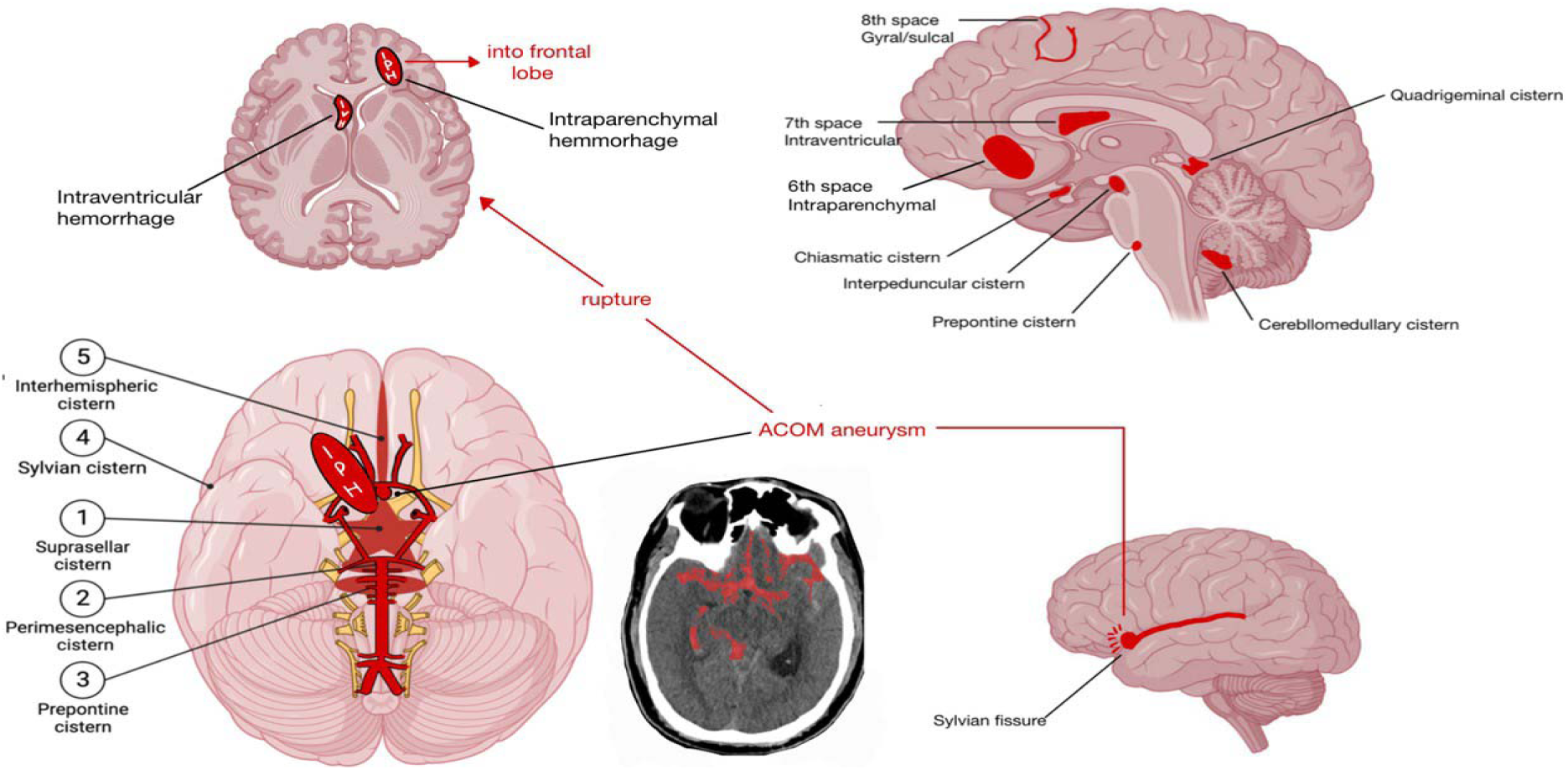
aSAH major intracranial cisternal blood locations Illustration depicting blood localization from different angles, specifically highlighting the most common rupture of the anterior communicating artery (Acom) into the frontal lobe, by BioRender.com

**Figure 6:**
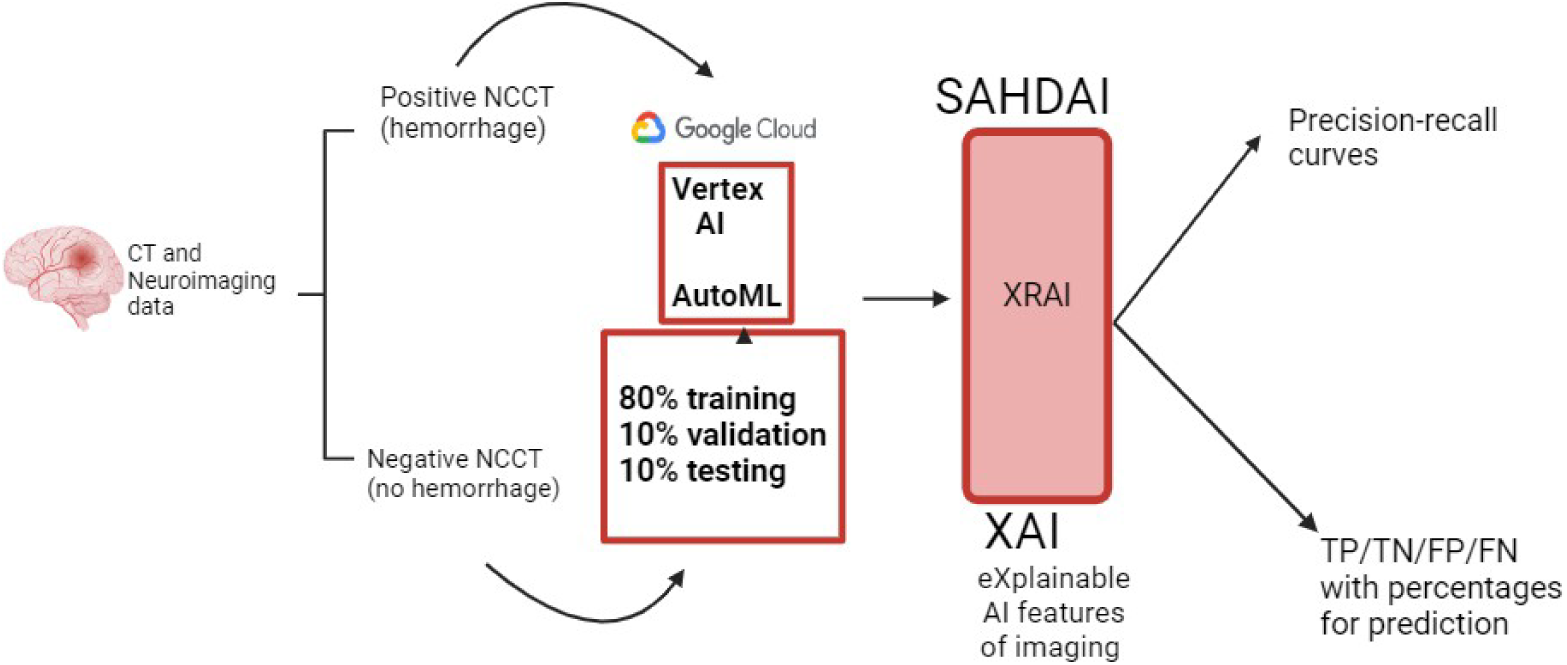
SAHDAI-XAI approach explanation, by biorender.com. SAHDAI- SAH Detection AI, XAI-eXplainable AI

**Figure 7:**
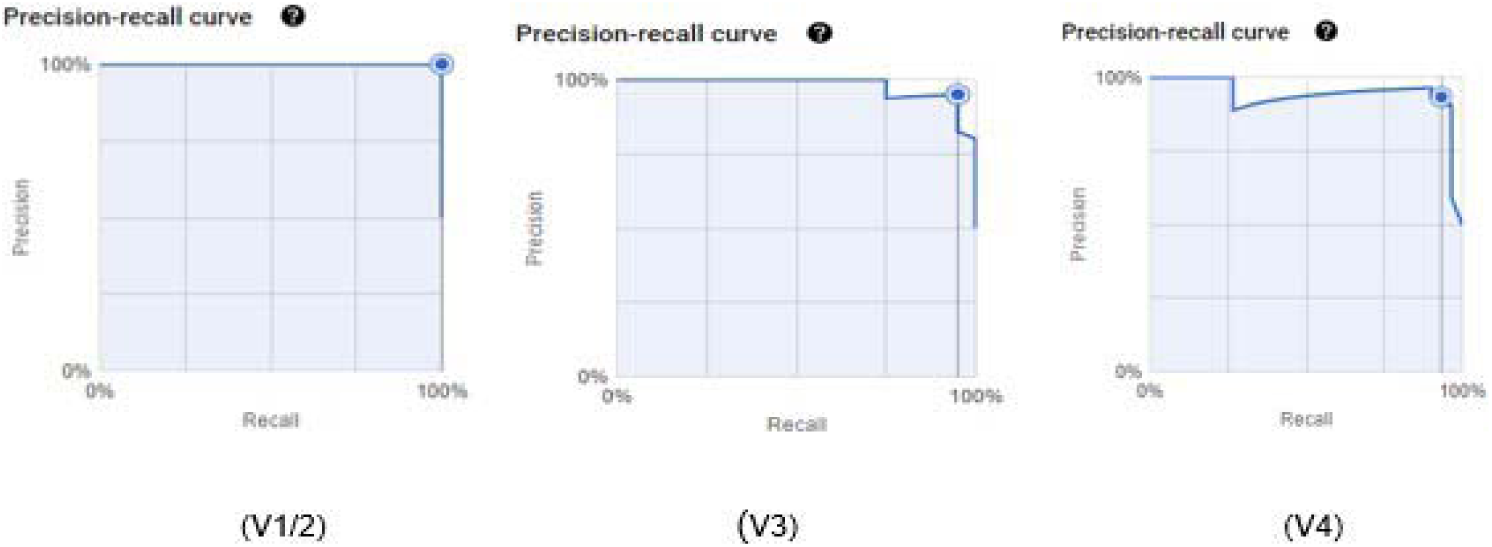
Precision-recall curves Precision-recall-curves on GCP for the four AutoML runs (V1-V4)

**Figure 8:**
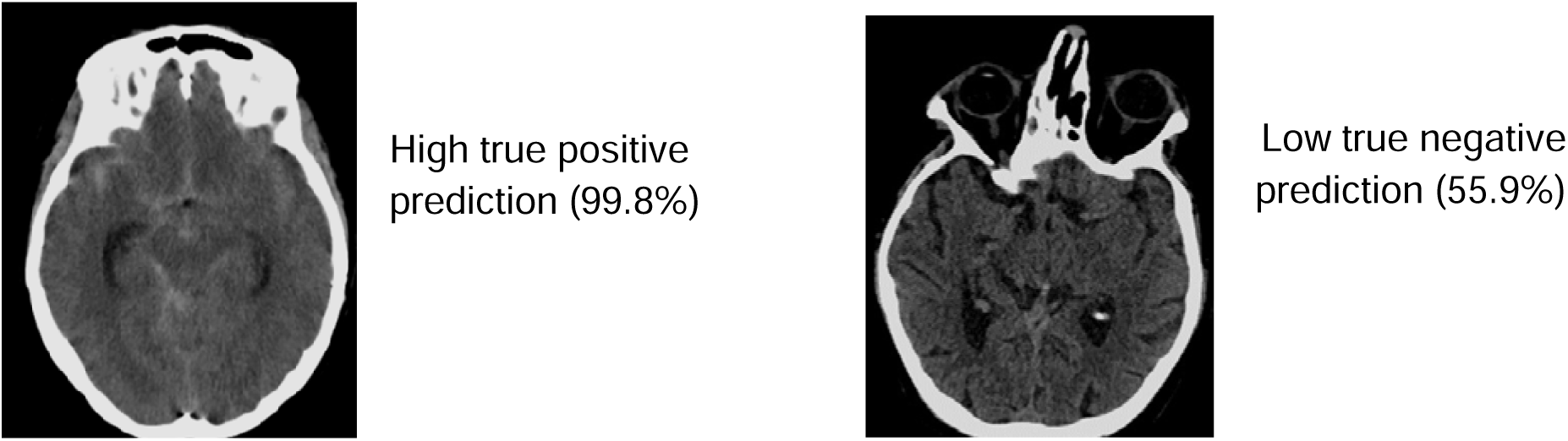
TP and TN examples illustration

**Figure 9:**
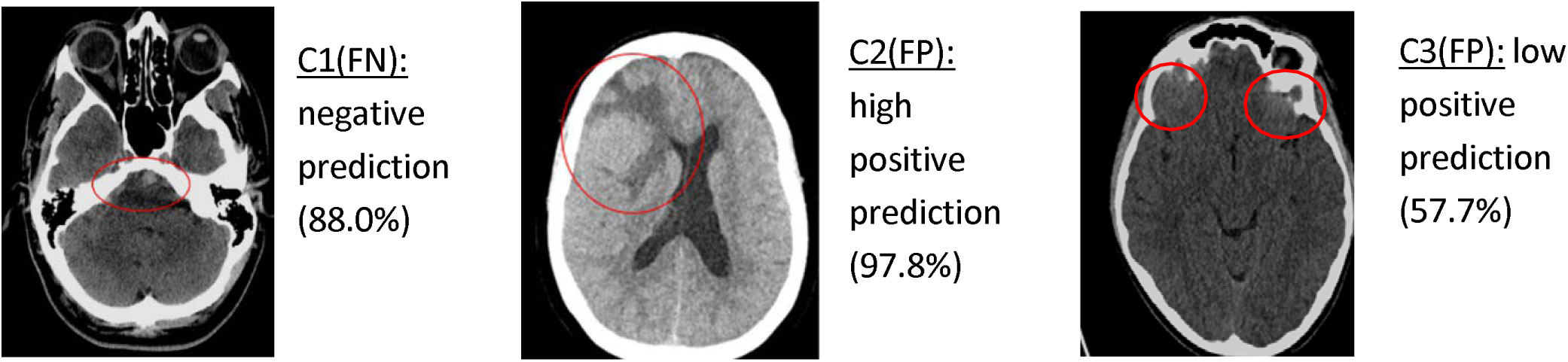
FP and FN predictions

**Figure 10:**
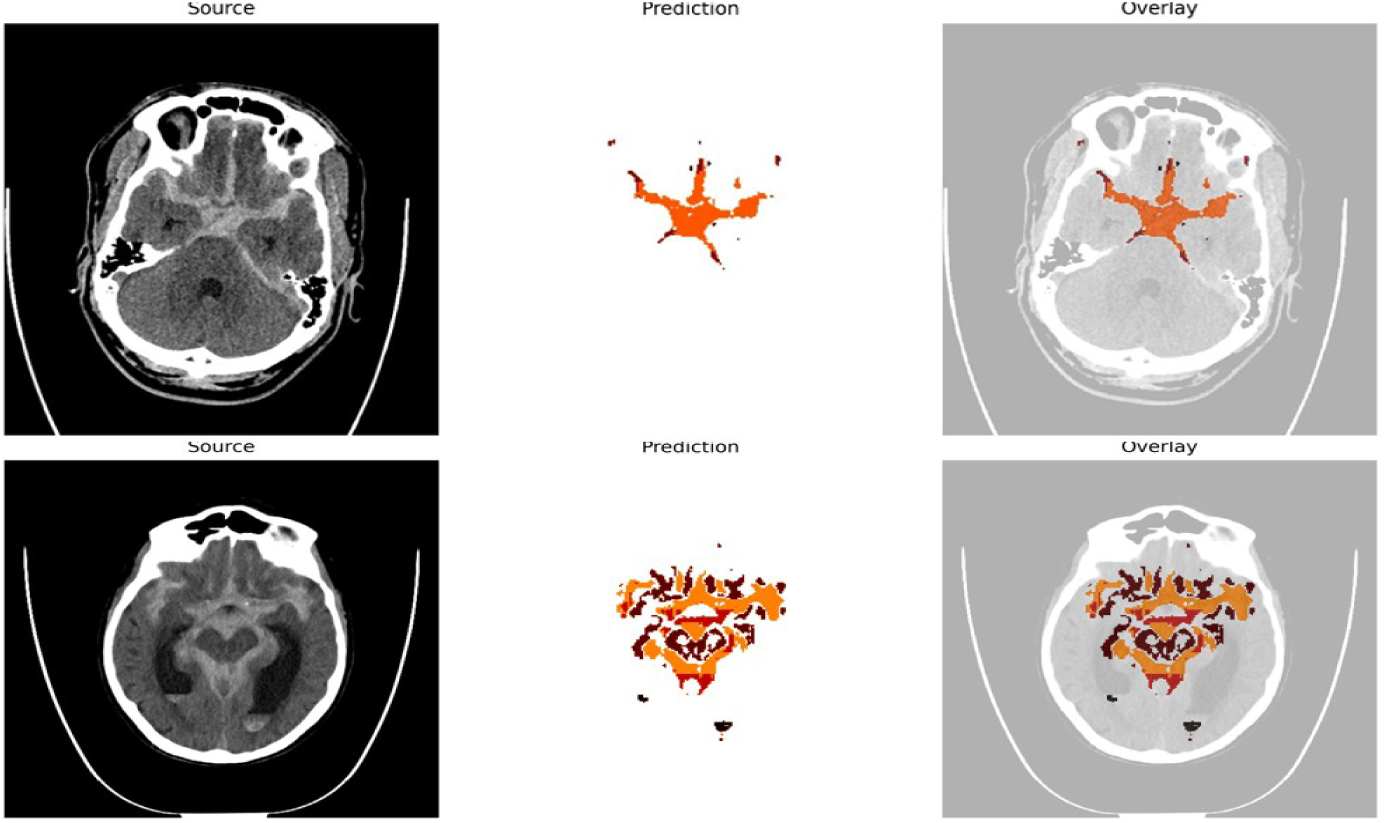
Category ***good***; above 60% color coverage of the hemorrhage

**Figure 11:**
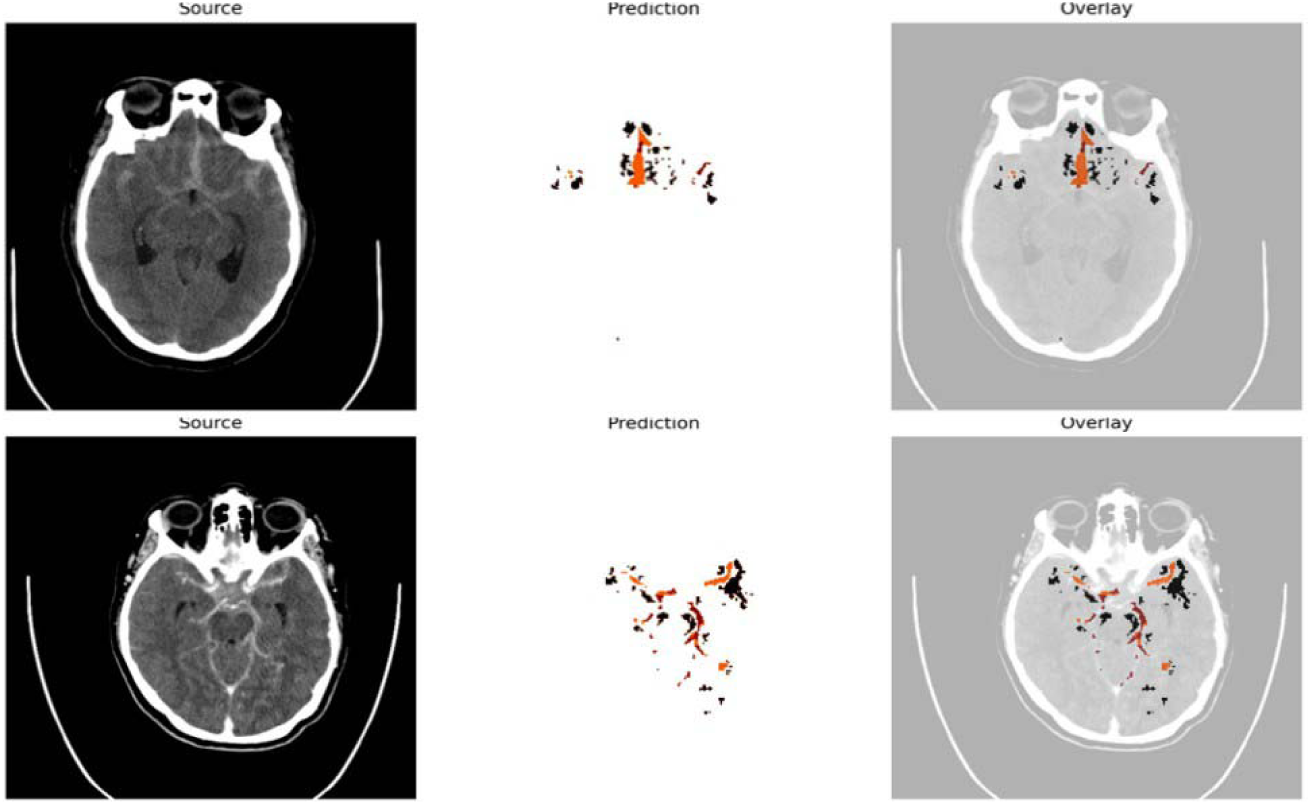
Category **moderate**; 30-60% color coverage of the hemorrhage

**Figure 12:**
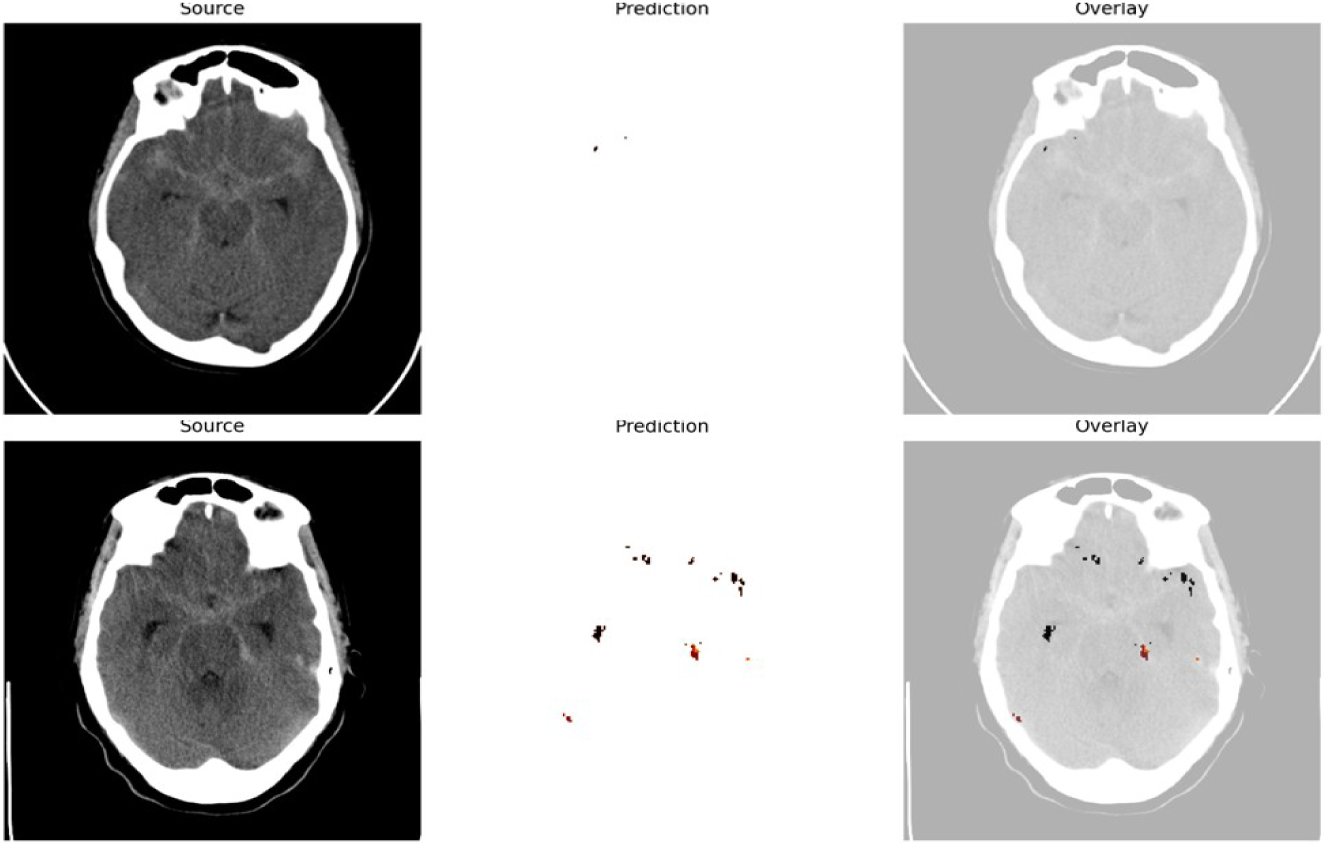
Category **bad**; less than 30% color coverage of the hemorrhage

**Figure 13:**
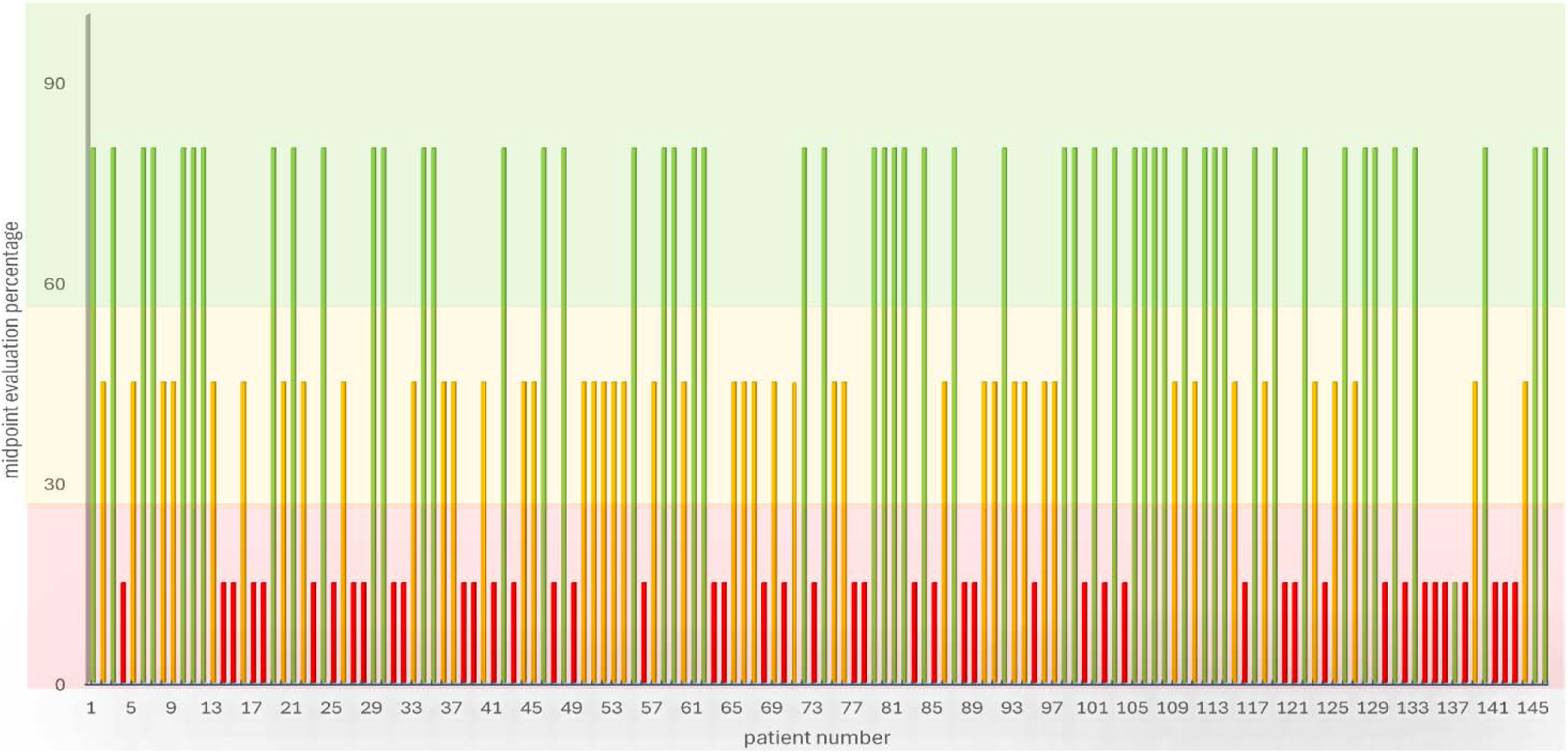
Descriptive evaluation of the BLAST-XAI model performance:

**Figure 14:**
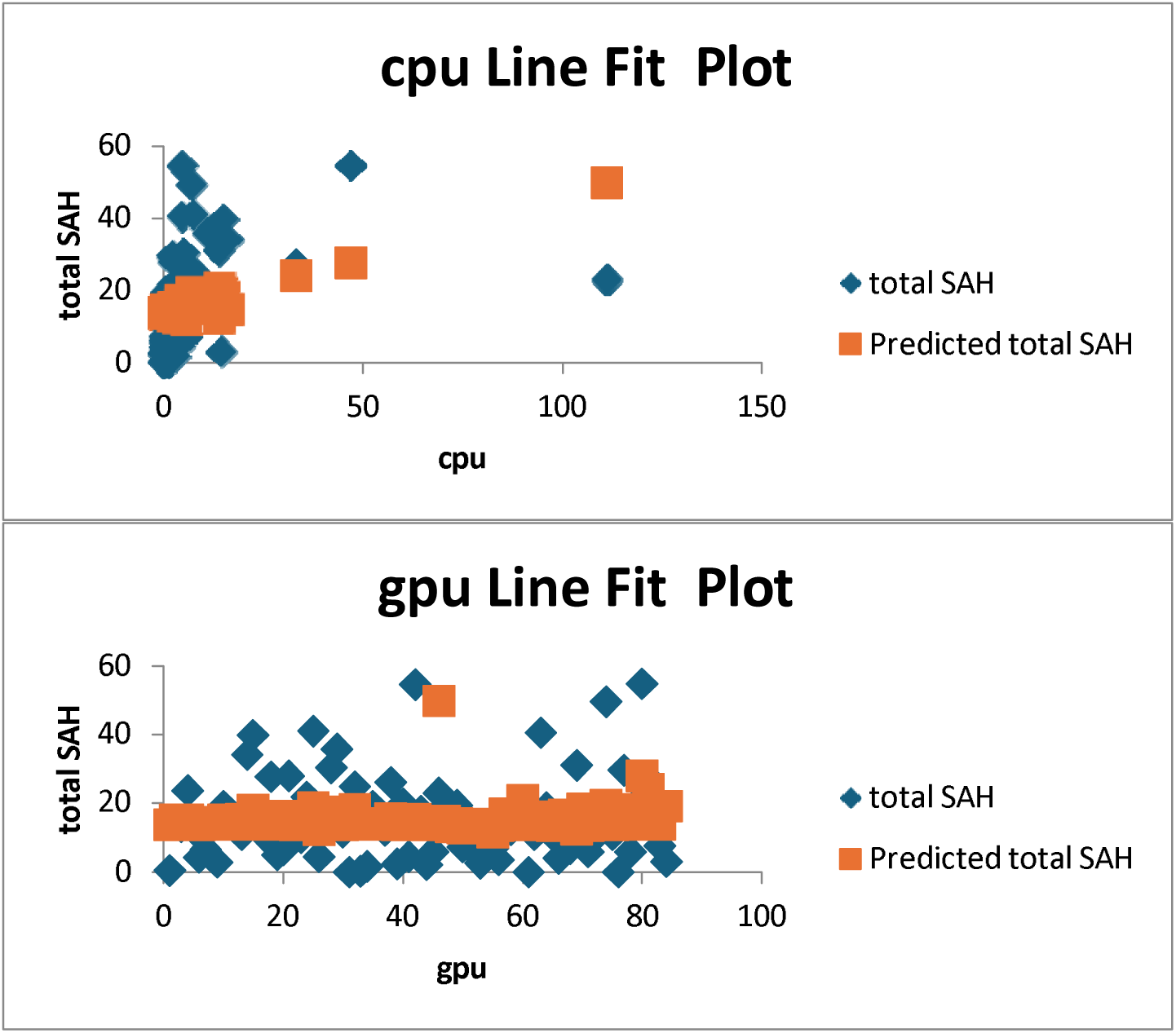
Graphical demonstration of the Correlation between BLAST-CT (CPU and GPU) predictions and ground truth SAHV in mL

**Figure 15:**
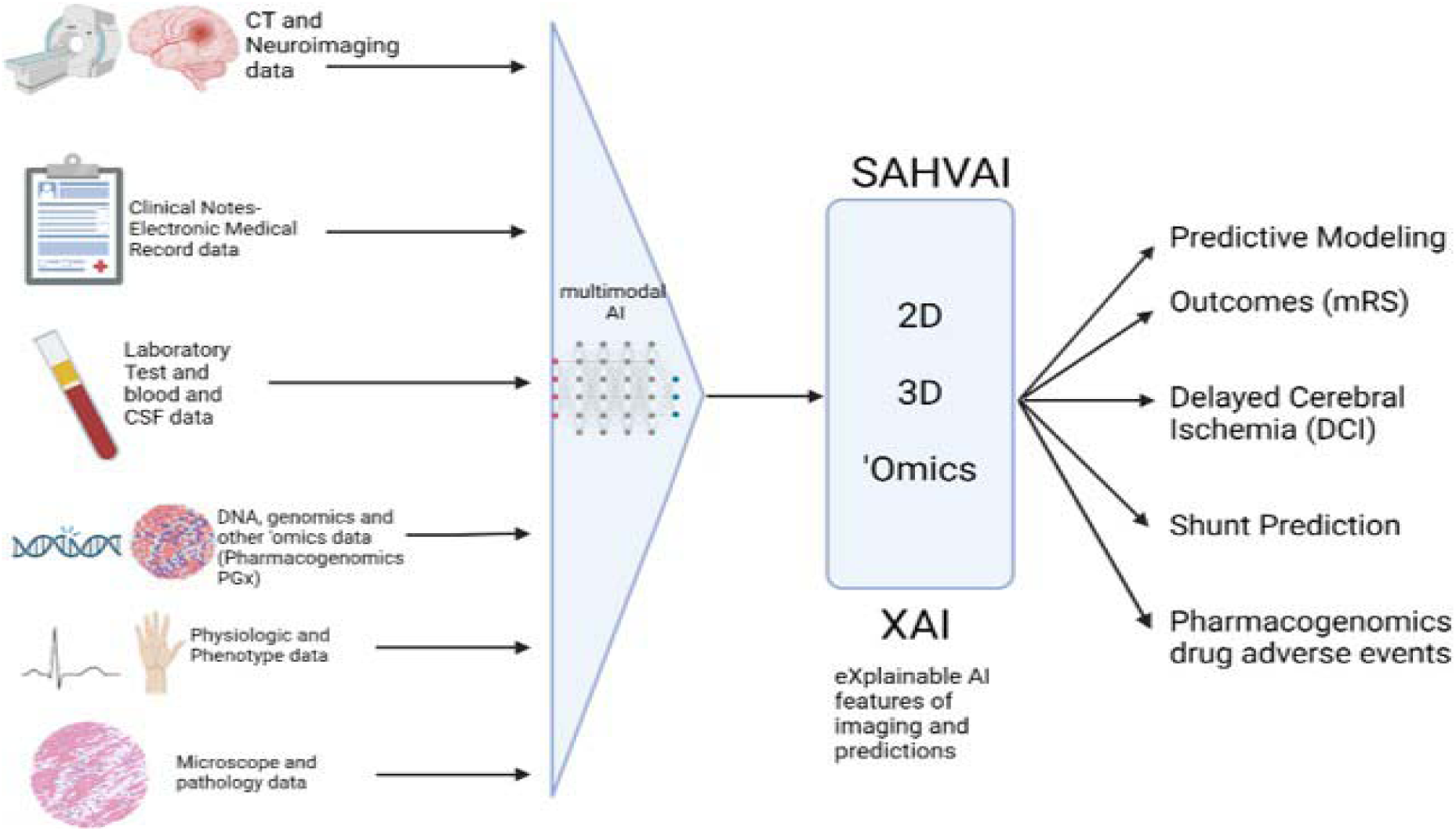
SAHVAI-Multi-modal model incorporates various inputs and includes SAHVAI-XAI for explainability of its predictions relative to actual outcomes

All in all, the R value demonstrated is relatively low but still statistically significant, which indicates a meaningful relationship between BLAST-CT predictions and the ground truth SAHV. The residuals are noticeably high, as shown in the ANOVA table. In some cases, the differences between AI predicted and manually measured blood volumes ranged from 7 to 15 mL. This level of variability lowers the R value and reduces the overall fit of the regression model. The model also tends to underestimate blood volume. However, when looking at the graphical results, a clear linear trend can be seen. As the hemorrhage volume increases, both BLAST-CT and manual measurements show similar trends. Furthermore, when the model was visually assessed, it became clear that the detection was more accurate in cases with larger bleeds, where the overlay masks covered a larger amount of the hemorrhage area. This supports the explainability of the SAHDAI-BLAST model both quantitatively and visually.

Finally, analyzing the BLAST-model performance and calculating the overlap measurements, the evaluation generated a Dice coefficient of 0.266 ± 0.21. In comparison, the SAHVAI model, which focuses on SAH blood volume segmentation and quantification, presented a dice score of 0.70 ± 0.06.

## 6 Discussion

A Stroke is a time-critical neurological emergency that demands an immediate medical response. It is the second leading cause of death globally, with ischemic stroke being the most prevalent type, followed by hemorrhagic stroke.^42,43^ SAH is a severe form of hemorrhagic stroke, typically due to an aneurysmal rupture. Even though it only represents 3% of all strokes, it carries a significant risk with high mortality rates.^44^ As a result of the dangerous and urgent nature of SAH, many AI medical models were developed to expedite the diagnosing process and save patients’ lives.^18,45^ However, to overcome AI’s untransparent black-box nature, it is essential to integrate explainability with the use of AI models in the medical field.^46^ The primary purpose of this study is to demonstrate the explainability of a cloud-based SAH detection model and to justify the predictions with visual predictive demonstrations.

### 6.1 Literature and Methodology Discussion

This study is split into two sequential phases. In the first phase, GCP’s Vertex AI was employed, aiming to develop a low-code AutoML binary classifying model to detect SAH and illustrate explainability.^47^ The low code aspect of AutoML can automate the machine learning process, creating the proper training and tuning environment. This approach simplifies it for medical professionals with no previous coding experience to develop AI models with no previous coding experience.^48^ Moreover, creating the SAHDAI-XAI model on GCP provides the advantage of presenting the model performance evaluation directly after training. This includes the display of precision-recall curves, average precision, and confusion matrices. Additionally, for each outcome, whether positive or negative, in this case, meaning hemorrhage or non-hemorrhage, the degree of prediction is expressed as a percentage, enhancing the explainability of the outcomes.

When Salman et al.^45^ created HEADS-UP on GCP, they aimed to develop a model to identify and evaluate intracranial hemorrhages (IH). However, the importance of XAI and visualizing the interpretations for AI predictions was highlighted after an FP outcome was presented in the study. The FP case demonstrated a left cerebellar IPH image, initially labeled as negative and later identified as hemorrhage by the model, without capturing the whole lesion. Inspired by the HEADS-UP study, a further enhancement of explainability with the main target of demonstrating reasons for a model’s predicted outcome transformed into the following priority of this study. This led to using the BLAST model’s publicly displayed code as a framework for the project’s second phase.^40^ The BLAST model has the combined ability to quantify brain images, for example, as presented in the study led by Chang et al., developing a neural network model that quantifies hemorrhages in head CT images^49^, and segmenting different regions as illustrated in the research conducted by Heit et al., using the software RAPID which is a neural network tool automating segmentation and detecting intracranial hemorrhages (ICH). This made the modal a more appealing choice to apply for SAH detection and explainability amplification. BLAST was initially designed for TBI detection, segmentation, and blood volume quantification for four lesion types, including IPH, EAH, edema, and IVH, without a clear focus on the SAH hemorrhage spaces. Therefore, the primary focus of SAH detection was on BLAST’s EAH identification with the consideration that SAH would be regarded as EAH.^36^

The exclusively descriptive BLAST versus Human approach aims to evaluate the model’s performance in its ability to detect SAH and visualize the explainability. A Python code adjustment was made to present parallel demonstrations of the source images, the predictions, and the masked overlay. This led to a better assessment of the detection ability, significantly increasing the model’s explainability.

Finally, the quantitative analysis of BLAST-CT using the SAHV variable as the manually calculated and measured ground truth, aimed to assess the model’s performance on both CPU and GPU configurations. Descriptive statistics and ANOVA regression were used to explore the modeĺs ability to detect SAH. The SAHV variable, based on expert measurements of CCT scans, proved to be reliable, easy to apply, and above all, explainable. These qualities make it a practical reference point for understanding model performance and considering its potential use in clinical settings.^12^

#### 6.1.2 Methodology limitations

One disadvantage presented with GCP usage is dependency. Without having access to the code, it becomes complicated to optimize and finetune the model in the case of errors or complications. Additionally, the dataset of CCTs is both randomized and anonymized, making it difficult to trace back specific clinical characteristics or demographic data for individual patients. This can make it challenging when trying to draw clinically important conclusions.

Another limitation is the categorization of the cases into good, moderate, or bad based on personal analysis as it could be perceived as subjective and may affect the objectivity and reproducibility of the findings.

Furthermore, the BLAST-CT model was originally developed for detecting TBI and not specifically designed to identify the eight potential bleeding spaces that are relevant in SAH detection. As a result, the model could have limitations to accurately identify all necessary bleeding compartments relevant to SAH, potentially reducing the diagnostic precision in this specific context.

### 6.2 Discussion of Results and potential clinical application

In Phase 1 of the study, the GCP Vertex AI AutoML SAHDAI-XAI model has an accurate and efficient performance with an average precision rate of 97.7% for SAH detection. After increasing the dataset size and challenging the model with other pathologies in the fourth AutoML run, the average precision rate decreased by 3.3% to 94.4%. These findings indicate that the model’s inability to differentiate between SAH hemorrhages and other hyperdense structures underlined the demonstration of two FP cases. However, it is essential to point out that the AI model detection of an FP outcome is considerably better than overseeing a time-critical SAH, considering the high mortality and morbidity rates of patients.^18,50^

After enhancing the visualization of explainability with the creation of the BLAST-SAHDAI-XAI model in the second phase of the model, its ability to detect SAH and demonstrate explainability has been proven. The calculated midpoint average of the descriptive performance assessment is 48.29%, reflecting a moderate model performance.

Adding a quantitative statistical analysis allowed for a more detailed assessment of the BLAST-CT model’s ability to detect SAH. Moreover, SAHV proved to be a reliable, measurable and explainable variable, making it a suitable ground truth reference.^12^ The model demonstrated statistical significance based on the p-value. However, the low regression value indicated that only a small proportion of the variance in the ground truth was explained by the model. Additionally, high residual variance and a large standard deviation of the prediction errors reflected notable variability in the model’s output. These statistical findings confirm the BLAST-CT model’s tendency to underestimate blood volume, which could affect its predictive performance and may limit its clinical implementation in its current form. It is necessary to further refine the model to improve its accuracy, reduce variability, and enhance its reliability for a routine clinical application. (Table 2-3.2)

Furthermore, a separate computed coefficient calculation comparing the BLAST model segmentation ability to label ground truth cases manually displayed a relatively low dice score of 0.266 ± 0.21. These results underline that the BLAST model was initially developed to segment and quantify TBI with different locations in the cranium based on the trauma and not for SAH.^36^ On the other hand, evaluating the performance of the SAHVAI model designed explicitly for aSAH segmentation and quantification showed a significantly better dice score of 0.70 ± 0.06, adding additional support to the aforementioned claim.These observed differences in performance could indicate that the BLAST-CT system may be working with random chance or coincidence, which raises questions about its potential clinical application, particularly when evaluating the difference in Dice scores between the SAHVAI model(0.7) and the SAHDAI-XAI-BLAST-CT model (0.226).

All in all, considering that this is the first attempt to focus on explainability for SAH detection based on qualitative imaging patterns, and given the lack of existing literature on this specific approach, the results can be seen as a promising starting point. However, since the BLAST-CT model was not specifically developed for the eight potential bleeding spaces associated with SAH, it can be seen that this affects the accuracy of its outputs. In daily clinical practice, the use of explainable AI in SAH detection would be essential to accelerate diagnosis and support more trustworthy clinical decision-making. However, at its current stage, the SAHDAI-XAI-BLAST-CT model is still not ready for clinical implementation and requires more refinement and code optimization specifically designed for SAH detection.

### 6.3 Future Directions

Although the results are promising, continued actions are required to optimize this approach and to achieve better outcomes, eventually expanding the project. To address the lack of differentiation between bleedings and hyperdense structures in the project’s first phase, more training with a dataset size expansion and running more AutoML rounds should be considered for better outcomes.^51^ Moreover, the moderate findings demonstrated in the second phase by the BLAST- SHDAIA-XAI model suggest that implementing another method specifically focusing on aSAH detection and explainability based on the eight potential hemorrhage spaces should be put into perspective. A better dice score indicating better segmentation performance with the SAHVAI model provides a possibility for future use of the SAHVAI model. This should target detection, blood volume quantification, and explainability.^19,36^

Furthermore, the long-term goal is to create a multimodal integrated model for SAH. Currently, the focus is on the detection and volumetric quantification. However, the vision is to develop a system integrating detection, quantification, past medical records, and other inputs with an additional integration of explainability. This is intended to generate trustworthy predictions, assisting healthcare professionals in accelerating diagnosis, supporting clinicians’ decisions, and ultimately saving patients’ lives.^52^

Finally, when looking for an effective and future-oriented method for the AI modelś usage enhancement in medicine, evaluative AI should be taken into consideration. This method aims to use decision-support tools to assist medical professionals in their decision-making with AI utilization. Instead of artificial intelligence just demonstrating the explanations it thinks are right, as observed in standard explainable AI, evaluative AI could help demonstrate more reliable results while still acknowledging the medical professionals’ suggestions and opinions. ^53^

## 7 Conclusion

SAHDAI-XAI and BLAST-SAHDAI-XAI are new image-based models presenting an advanced SAH detection method to illustrate explainability. These models facilitate a fast-diagnostic method for SAH, representing a neurological time-critical disease, with the help of explainability factors integration. This strives to demonstrate and enhance the comprehensibility of predicted outcomes by overcoming the untransparent black-box nature of AI. Focusing on the prospective improvement of interpretability and trustworthiness for SAH detection, with the intention of assisting medical professionals in an expedited, accurate, and explainable medical evaluation, and ultimately improving the patient standard of care and lowering mortality and morbidity rates, is this project’s main objective. However, the model still requires further refinement, as it still not capable of reliably detecting all areas of SAH bleeding in a CCT scan.

## Data Availability

All data produced in the present study are available upon reasonable request to the authors

## References

1. Long B, Koyfman A, Runyon MS. Subarachnoid Hemorrhage: Updates in Diagnosis and Management. Emerg Med Clin North Am. Nov 2017;35(4):803–824. doi:10.1016/j.emc.2017.07.001

2. Hoh BL, Ko NU, Amin-Hanjani S, et al. 2023 Guideline for the Management of Patients With Aneurysmal Subarachnoid Hemorrhage: A Guideline From the American Heart Association/American Stroke Association. Stroke. Jul 2023;54(7):e314-e370. doi:10.1161/str.0000000000000436

3. Petridis AK, Kamp MA, Cornelius JF, et al. Aneurysmal Subarachnoid Hemorrhage. Dtsch Arztebl International. March 31, 2017 2017;114(13):226-36. doi:10.3238/arztebl.2017.0226

4. Francoeur CL, Mayer SA. Management of delayed cerebral ischemia after subarachnoid hemorrhage. Critical Care. 2016/10/14 2016;20(1):277. doi:10.1186/s13054-016-1447-6

5. O’Carroll CB, Brown BL, Freeman WD. Intracerebral Hemorrhage: A Common yet Disproportionately Deadly Stroke Subtype. Mayo Clin Proc. Jun 2021;96(6):1639–1654. doi:10.1016/j.mayocp.2020.10.034

6. Marcolini E, Hine J. Approach to the Diagnosis and Management of Subarachnoid Hemorrhage. West J Emerg Med. Mar 2019;20(2):203–211. doi:10.5811/westjem.2019.1.37352

7. Liu J, Sun C, Wang Y, et al. Efficacy of nimodipine in the treatment of subarachnoid hemorrhage: a meta-analysis. Arq Neuropsiquiatr. Jul 2022;80(7):663–670. Eficácia da nimodipina no tratamento da hemorragia subaracnoidea: uma metanálise. doi:10.1055/s-0042-1755301

8. Naidech AM. Intracranial hemorrhage. Am J Respir Crit Care Med. Nov 1 2011;184(9):998–1006. doi:10.1164/rccm.201103-0475CI

9. Park S-H, Kim TJ, Ko S-B. Transcranial Doppler Monitoring in Subarachnoid Hemorrhage. J Neurosonol Neuroimag. 6 2022;14(1):1–9. doi:10.31728/jnn.2022.00115

10. Tawk RG, Hasan TF, D’Souza CE, Peel JB, Freeman WD. Diagnosis and Treatment of Unruptured Intracranial Aneurysms and Aneurysmal Subarachnoid Hemorrhage. Mayo Clin Proc. Jul 2021;96(7):1970–2000. doi:10.1016/j.mayocp.2021.01.005

11. Lagares A, Jiménez-Roldán L, Gomez PA, et al. Prognostic Value of the Amount of Bleeding After Aneurysmal Subarachnoid Hemorrhage: A Quantitative Volumetric Study. Neurosurgery. 2015;77(6):898–907. doi:10.1227/neu.0000000000000927

12. Foettinger F, Sharma R, Salman S, et al. The ABCs of Subarachnoid Hemorrhage Blood Volume Measurement: A Simplified Quantitative Method Predicts Outcomes and Delayed Cerebral Ischemia. medRxiv. 2023:2023.09.05.23295090. doi:10.1101/2023.09.05.23295090

13. Sharma R, Mandl D, Foettinger F, et al. The eSAH Score: A Simple Practical Predictive Model for SAH Mortality & Outcomes. medRxiv. 2023:2023.09.15.23295634. doi:10.1101/2023.09.15.23295634

14. Shi Z, Hu B, Schoepf UJ, et al. Artificial Intelligence in the Management of Intracranial Aneurysms: Current Status and Future Perspectives. AJNR Am J Neuroradiol. Mar 2020;41(3):373–379. doi:10.3174/ajnr.A6468

15. Silva MA, Patel J, Kavouridis V, et al. Machine Learning Models can Detect Aneurysm Rupture and Identify Clinical Features Associated with Rupture. World Neurosurg. Nov 2019;131:e46–e51. doi:10.1016/j.wneu.2019.06.231

16. Zhang B, Shi H, Wang H. Machine Learning and AI in Cancer Prognosis, Prediction, and Treatment Selection: A Critical Approach. J Multidiscip Healthc. 2023;16:1779–1791. doi:10.2147/jmdh.S410301

17. Alowais SA, Alghamdi SS, Alsuhebany N, et al. Revolutionizing healthcare: the role of artificial intelligence in clinical practice. BMC Medical Education. 2023/09/22 2023;23(1):689. doi:10.1186/s12909-023-04698-z

18. Salman S, Gu Q, Sharma R, et al. Artificial intelligence and machine learning in aneurysmal subarachnoid hemorrhage: Future promises, perils, and practicalities. Journal of the Neurological Sciences. 2023/11/15/ 2023;454:120832. 10.1016/j.jns.2023.120832

19. Wirtz M, Salman S, Wei Y, et al. SAHVAI-3D and 4D: A New, Automated Subarachnoid Hemorrhage Volumetric Artificial Intelligence (SAHVAI) Measurement Approach Using Non-Contrast Head CT Scans. 2024:

20. Barredo Arrieta A, Díaz-Rodríguez N, Del Ser J, et al. Explainable Artificial Intelligence (XAI): Concepts, taxonomies, opportunities, and challenges toward responsible AI. Information Fusion. 2020/06/01/ 2020;58:82-115. 10.1016/j.inffus.2019.12.012

21. van der Velden BHM, Kuijf HJ, Gilhuijs KGA, Viergever MA. Explainable artificial intelligence (XAI) in deep learning-based medical image analysis. Med Image Anal. Jul 2022;79:102470. doi:10.1016/j.media.2022.102470

22. Department of Health and Human Services CfMMS, Office of the Secretary *Nondiscrimination in Health Programs and Activities*. Vol. 89. Rule. 2024:37522-37703 0945-AA17. May 6, 2024. https://www.govinfo.gov/content/pkg/FR-2024-05-06/pdf/2024-08711.pdf

23. Saeed W, Omlin C. Explainable AI (XAI): A systematic meta-survey of current challenges and future opportunities. Knowledge-Based Systems. 2023/03/05/ 2023;263:110273. 10.1016/j.knosys.2023.110273

24. Nazir S, Dickson DM, Akram MU. Survey of explainable artificial intelligence techniques for biomedical imaging with deep neural networks. Computers in Biology and Medicine. 2023/04/01/ 2023;156:106668. 10.1016/j.compbiomed.2023.106668

25. Islam SR, Eberle W, Ghafoor SK, Ahmed M. Explainable Artificial Intelligence Approaches: A Survey. ArXiv. 2021;abs/2101.09429

26. Holzinger A, Saranti A, Molnar C, Biecek P, Samek W. Explainable AI Methods - A Brief Overview. In: Holzinger A, Goebel R, Fong R, Moon T, Müller K-R, Samek W, eds. xxAI - Beyond Explainable AI: International Workshop, Held in Conjunction with ICML 2020*, July 18, 2020, Vienna, Austria, Revised and Extended Papers*. Springer International Publishing; 2022:13-38.

27. Kapishnikov AB, Tolga; Viegas, Fernanda; Terry, Michael. XRAI: Better Attributions Through Regions. 2019:4947–4956. doi:10.1109/ICCV.2019.00505

28. Avram MG. Advantages and Challenges of Adopting Cloud Computing from an Enterprise Perspective. Procedia Technology. 2014/01/01/ 2014;12:529-534. 10.1016/j.protcy.2013.12.525

29. Rokis K, Kirikova M. Exploring Low-Code Development: A Comprehensive Literature Review. Complex Systems Informatics and Modeling Quarterly. 10/31 2023;0:68–86. doi:10.7250/csimq.2023-36.04

30. Thiriveedhi VK, Krishnaswamy D, Clunie D, Pieper S, Kikinis R, Fedorov A. Cloud-based large-scale curation of medical imaging data using AI segmentation. Res Sq. May 3 2024;doi:10.21203/rs.3.rs-4351526/v1

31. Sachdeva S, Bhatia S, Al Harrasi A, et al. Unraveling the role of cloud computing in health care system and biomedical sciences. Heliyon. Apr 15 2024;10(7):e29044. doi:10.1016/j.heliyon.2024.e29044

32. Challita S ZF, Gourdin C, Merle P. A Precise Model for Google Cloud Platform. presented at: 2018 IEEE International Conference on Cloud Engineering (IC2E); 2018; Orlando, FL, USA.

33. Salehin I, Islam MS, Saha P, et al. AutoML: A systematic review on automated machine learning with neural architecture search. Journal of Information and Intelligence. 2024/01/01/ 2024;2(1):52-81. 10.1016/j.jiixd.2023.10.002

34. He X, Zhao K, Chu X. AutoML: A survey of the state-of-the-art. Knowledge-Based Systems. 2021/01/05/ 2021;212:106622. 10.1016/j.knosys.2020.106622

35. Heit JJ, Coelho H, Lima FO, et al. Automated Cerebral Hemorrhage Detection Using RAPID. American Journal of Neuroradiology. 2021;42(2):273. doi:10.3174/ajnr.A6926

36. Monteiro M, Newcombe VFJ, Mathieu F, et al. Multiclass semantic segmentation and quantification of traumatic brain injury lesions on head CT using deep learning: an algorithm development and multicentre validation study. The Lancet Digital Health. 2020;2(6):e314–e322. doi:10.1016/S2589-7500(20)30085-6

37. Yushkevich PA, Yang G, Gerig G. ITK-SNAP: An interactive tool for semi-automatic segmentation of multi-modality biomedical images. Annu Int Conf IEEE Eng Med Biol Soc. Aug 2016;2016:3342–3345. doi:10.1109/embc.2016.7591443

38. Cloud G. Explainable AI Overview. Accessed September 20, 2024, https://cloud.google.com/vertex-ai/docs/explainable-ai/overview

39. Fortner B MT. Number by Colors: A Guide To Using Color To Understand Technical Data. 1 ed. Springer 1997:1–349.

40. biomedia-mira. BLAST-CT [software]. Accessed October 1, 2024, https://github.com/biomedia-mira/blast-ct

41. Eelbode T, Bertels J, Berman M, et al. Optimization for Medical Image Segmentation: Theory and Practice When Evaluating With Dice Score or Jaccard Index. IEEE Trans Med Imaging. Nov 2020;39(11):3679–3690. doi:10.1109/tmi.2020.3002417

42. Saver JL. Time Is Brain—Quantified. Stroke. 2006/01/01 2006;37(1):263–266. doi:10.1161/01.STR.0000196957.55928.ab

43. Feigin VL, Brainin M, Norrving B, et al. World Stroke Organization (WSO): Global Stroke Fact Sheet 2022. Int J Stroke. Jan 2022;17(1):18–29. doi:10.1177/17474930211065917

44. Muehlschlegel S. Subarachnoid Hemorrhage. *Continuum (Minneap Minn*). Dec 2018;24(6):1623-1657. doi:10.1212/con.0000000000000679

45. Salman S, Gu Q, Dherin B, et al. Hemorrhage Evaluation and Detector System for Underserved Populations: HEADS-UP. Mayo Clinic Proceedings: Digital Health. 2023/12/01/ 2023;1(4):547–556. 10.1016/j.mcpdig.2023.08.009

46. Doshi-Velez F, Kim B. Towards A Rigorous Science of Interpretable Machine Learning. arXiv: Machine Learning. 2017;

47. Bisong E. An Overview of Google Cloud Platform Services. Building Machine Learning and Deep Learning Models on Google Cloud Platform: A Comprehensive Guide for Beginners. Apress; 2019:7–10.

48. Shin S, Park D, Ji S, Joo G, Im H. Medical Data Analysis Using AutoML Frameworks. Journal of Electrical Engineering & Technology. 2024/09/01 2024;19(7):4515-4522. doi:10.1007/s42835-024-01919-3

49. Chang PD, Kuoy E, Grinband J, et al. Hybrid 3D/2D Convolutional Neural Network for Hemorrhage Evaluation on Head CT. AJNR Am J Neuroradiol. Sep 2018;39(9):1609–1616. doi:10.3174/ajnr.A5742

50. Edlow JA. Diagnosis of subarachnoid hemorrhage. Neurocrit Care. 2005;2(2):99–109. doi:10.1385/ncc:2:2:099

51. Uddin S, Lu H. Dataset meta-level and statistical features affect machine learning performance. Scientific Reports. 2024/01/19 2024;14(1):1670. doi:10.1038/s41598-024-51825-x

52. Pahud de Mortanges A, Luo H, Shu SZ, et al. Orchestrating explainable artificial intelligence for multimodal and longitudinal data in medical imaging. npj Digital Medicine. 2024/07/22 2024;7(1):195. doi:10.1038/s41746-024-01190-w

53. Miller T. Explainable AI is dead, long live explainable AI! Hypothesis-driven decision support using evaluative AI. Proc 6th ACM Conf Fairness Accountability Transpar (FAccT). 2023:3593013–3594001. doi:10.1145/3593013.3594001

54. Hillis, James M, et al. “Evaluation of an Artificial Intelligence Model for Identification of Intracranial Hemorrhage Subtypes on Computed Tomography of the Head.” Stroke Vascular and Interventional Neurology, vol. 4, no. 4, 16 May 2024, 10.1161/svin.123.001223.

55. Shu L, Yan H, Wu Y, et al. Explainable machine learning in outcome prediction of high-grade aneurysmal subarachnoid hemorrhage. Aging (Albany NY). 2024;16(5):4654–4669. doi:10.18632/aging.205621

56. Mohammadzadeh I, Niroomand B, Shahnazian Z, et al. Machine learning for predicting poor outcomes in aneurysmal subarachnoid hemorrhage: A systematic review and meta-analysis involving 8445 participants. Clinical Neurology and Neurosurgery. 2024;249:108668. 10.1016/j.clineuro.2024.108668

57. Kamdar N. Text Classification using AutoML Tables|Google Cloud Platform. Medium. Published February 15, 2022. Accessed April 17, 2025. https://medium.com/google-cloud/text-classification-using-automl-tables-google-cloud-platform-43816c4afb93

58. García-García S, Cepeda S, Dominik Müller, et al. Mortality Prediction of Patients with Subarachnoid Hemorrhage Using a Deep Learning Model Based on an Initial Brain CT Scan. Brain Sciences. 2023;14(1):10–10. 10.3390/brainsci14010010

59. Zarrin DA, Suri A, McCarthy K, et al. Machine learning predicts cerebral vasospasm in patients with subarachnoid haemorrhage. EBioMedicine. 2024;105:105206–105206. 10.1016/j.ebiom.2024.105206

60. Kim KH, Koo HW, Lee BJ, Sohn MJ. Analysis of risk factors correlated with angiographic vasospasm in patients with aneurysmal subarachnoid hemorrhage using explainable predictive modeling. Journal of Clinical Neuroscience. 2021;91:334–342. 10.1016/j.jocn.2021.07.028

61. Gombolay GY, Silva A, Schrum M, et al. Effects of explainable artificial intelligence in neurology decision support. Annals of Clinical and Translational Neurology. 2024;11(5):1224–1235. 10.1002/acn3.52036

62. Srivastava P. CPU vs GPU: Architectural Differences, Performance Metrics, and Use Cases. Medium. Published February 21, 2025. Accessed April 19, 2025. https://medium.com/%40prabhuss73/cpu-vs-gpu-architectural-differences-performance-metrics-and-use

